# Ethnic differences in SARS-CoV-2 vaccine hesitancy in United Kingdom healthcare workers: Results from the UK-REACH prospective nationwide cohort study

**DOI:** 10.1101/2021.04.26.21255788

**Authors:** Katherine Woolf, I Chris McManus, Christopher A Martin, Laura B Nellums, Anna L Guyatt, Carl Melbourne, Luke Bryant, Mayuri Gogoi, Fatimah Wobi, Amani Al-Oraibi, Osama Hassan, Amit Gupta, Catherine John, Martin D Tobin, Sue Carr, Sandra Simpson, Bindu Gregary, Avinash Aujayeb, Stephen Zingwe, Rubina Reza, Laura J Gray, Kamlesh Khunti, Manish Pareek, On behalf of the UK-REACH Study Collaborative Group

## Abstract

**Background:** In most countries, healthcare workers (HCWs) represent a priority group for vaccination against severe acute respiratory syndrome coronavirus-2 (SARS-CoV-2) due to their elevated risk of COVID-19 and potential contribution to nosocomial SARS-CoV-2 transmission. Concerns have been raised that HCWs from ethnic minority groups are more likely to be vaccine hesitant (defined by the World Health Organisation as refusing or delaying a vaccination) than those of White ethnicity, but there are limited data on SARS-CoV-2 vaccine hesitancy and its predictors in UK HCWs.

**Methods:** Nationwide prospective cohort study and qualitative study in a multi-ethnic cohort of clinical and non-clinical UK HCWs. We analysed ethnic differences in SARS-CoV-2 vaccine hesitancy adjusting for demographics, vaccine trust, and perceived risk of COVID-19. We explored reasons for hesitancy in qualitative data using a framework analysis.

**Findings:** 11,584 HCWs were included in the cohort analysis. 23% (2704) reported vaccine hesitancy. Compared to White British HCWs (21.3% hesitant), HCWs from Black Caribbean (54.2%), Mixed White and Black Caribbean (38.1%), Black African (34.4%), Chinese (33.1%), Pakistani (30.4%), and White Other (28.7%) ethnic groups were significantly more likely to be hesitant. In adjusted analysis, Black Caribbean (aOR 3.37, 95% CI 2.11 - 5.37), Black African (aOR 2.05, 95% CI 1.49 - 2.82), White Other ethnic groups (aOR 1.48, 95% CI 1.19 - 1.84) were significantly more likely to be hesitant. Other independent predictors of hesitancy were younger age, female sex, higher score on a COVID-19 conspiracy beliefs scale, lower trust in employer, lack of influenza vaccine uptake in the previous season, previous COVID-19, and pregnancy. Qualitative data from 99 participants identified the following contributors to hesitancy: lack of trust in government and employers, safety concerns due to the speed of vaccine development, lack of ethnic diversity in vaccine studies, and confusing and conflicting information. Participants felt uptake in ethnic minority communities might be improved through inclusive communication, involving HCWs in the vaccine rollout, and promoting vaccination through trusted networks.

**Interpretation:** Despite increased risk of COVID-19, HCWs from some ethnic minority groups are more likely to be vaccine hesitant than their White British colleagues. Strategies to build trust and dispel myths surrounding the COVID-19 vaccine in these communities are urgently required. Public health communications should be inclusive, non-stigmatising and utilise trusted networks.

**Funding:** MRC-UK Research and Innovation (MR/V027549/1), the Department of Health and Social Care through the National Institute for Health Research (NIHR), and NIHR Biomedical Research Centres and NIHR Applied Research Collaboration East Midlands.

**Research in context:** *Evidence before this study:* We searched Pubmed using the following search terms ((COVID-19).ti,ab OR (SARS-CoV-2).ti,ab) AND ((vaccine).ti,ab OR (vaccination).ti,ab OR (immunisation).ti,ab)) AND ((healthcare worker).ti,ab OR (health worker).ti,ab OR (doctor).ti,ab OR (nurse).ti,ab OR (healthcare professional).ti,ab)) AND ((hesitancy).ti,ab OR (refusal).ti,ab OR (uptake).ti,ab)). The search returned 60 results, of which 38 were excluded after title and abstract screening, 11 studies were not conducted in a population of healthcare workers, 20 did not present data on vaccine intention or uptake, 5 were related to vaccines other than the SARS-CoV-2 vaccine, 1 was unrelated to vaccination and 1 had been withdrawn. The 22 remaining articles were survey studies focussed on SARS-CoV-2 vaccine intention in healthcare workers. Estimates of SARS-CoV-2 vaccine acceptance varied widely from 27·7% - 94·5% depending on the country in which the study was performed, and the occupational group studied. Only 2 studies (both conducted in the USA) had a sample size greater than 10,000. Most studies found females, non-medical healthcare staff and those refusing influenza vaccine to be more likely to be hesitant. There was conflicting evidence about the effects of age and previous COVID-19 on hesitancy. Only 3 studies (all from the USA), presented data disaggregated by ethnicity, all finding Black ethnic HCWs were most likely to be hesitant. Common themes amongst studies that investigated reasons for vaccine hesitancy were concerns about safety of vaccines, fear of side effects and short development timeframes. We did not find any studies on SARS-CoV-2 vaccine hesitancy in UK healthcare workers in the published literature.

*Added value of this study:* This study is amongst the largest SARS-CoV-2 vaccine hesitancy studies in the literature. It is the largest study outside the USA and is the only study in UK HCWs. Our work focusses on the association of ethnicity with vaccine hesitancy, and we are the first study outside the USA to present results by ethnic group. The large number of ethnic minority HCWs in our study allows for examination of the outcome by more granular ethnicity categories than have previously been studied, allowing us to detect important differences in vaccine hesitancy levels within the broad White and Asian ethnic groupings. Our large sample size and the richness of our cohort study dataset allows us to control for many potential confounders in our multivariable analysis, and provide novel data on important potential drivers of hesitancy including discrimination, COVID-19 conspiracy beliefs, religion/religiosity and personality traits. Additionally, we combine quantitative with qualitative data providing a deeper understanding of the drivers of hesitancy and potential strategies to improve vaccine uptake in HCWs from ethnic minority communities.

*Implications of all the available evidence:* Around a quarter of UK healthcare workers reported SARS-CoV-2 vaccine hesitancy. In accordance with previous studies in other countries, we determined that female sex and lack of influenza vaccine in the previous season were important predictors of SARS-CoV-2 vaccine hesitancy in UK HCWs, although in contrast to most studies in the published literature, after adjustment we do not demonstrate differences in hesitancy levels by occupational role. Importantly, previous literature provides conflicting evidence of the effects of age and previous SARS-CoV-2 infection on vaccine hesitancy. In our study, younger HCWs and those with evidence of previous COVID-19 were more likely to be hesitant. This study provides novel data on increased hesitancy levels within Black Caribbean, Mixed White and Black Caribbean, Black African, Chinese, Pakistani and White Other ethnic groups. Mistrust (of vaccines in general, in SARS-CoV-2 vaccines specifically, in healthcare systems and research) and misinformation appear to be important drivers of hesitancy within HCWS in the UK. Our data indicate that despite facing an increased risk of COVID-19 compared to their White colleagues, UK HCWs from some ethnic minority groups continue to exhibit greater levels of SARS-CoV-2 vaccine hesitancy. This study provides policy makers with evidence to inform strategies to improve uptake.

## Introduction

An unprecedented global research effort has resulted in effective vaccines against the causative agent of COVID-19, severe acute respiratory syndrome coronavirus 2 (SARS-CoV-2).^1,2^ Emerging evidence suggests that mass vaccination programmes, which are underway globally, can significantly reduce the incidence of COVID-19 infections, hospitalisations and deaths.^3^ The UK Joint Committee on Vaccination and Immunisation (JCVI) have prioritised certain high-risk groups in the UK’s vaccination programme, including frontline health and social care staff. There are however concerns about SARS-CoV-2 vaccine hesitancy among healthcare workers (HCWs),^4–13^ and particularly among some ethnic minority groups ^14–21^ including ethnic minority HCWs ^22,23^ despite those groups being disproportionately affected by the pandemic.^24,25^

Vaccine hesitancy is defined by the World Health Organisation (WHO) as refusal or delay in vaccine acceptance.^26^ Levels of hesitancy towards specific vaccines and/or vaccines in general differ across individuals. Vaccine hesitancy amongst HCWs is especially concerning because it increases risks to the health of the individual HCW, is likely to increase the risk of nosocomial SARS-CoV-2 transmission,^27^ and may influence patient vaccine uptake.^15,28^ Reasons for vaccine hesitancy vary between individuals and by context, geographic location and vaccine, but the WHO’s “Three C’s model” has identified three areas influencing hesitancy: Convenience (vaccine access), Confidence (trust - in vaccines generally, in their efficacy, in those providing the vaccine, and in those creating vaccine policy), and Complacency (perceived risk of vaccine-related disease).^26,28–30^

The UK’s Scientific Advisory Group for Emergencies (SAGE) ethnicity subgroup has identified the following key factors underlying vaccine hesitancy in ethnic minority groups: physical barriers to access; lower trust and confidence in vaccine efficacy and safety, and general lack of trust in healthcare and health research due to structural and institutional racism and discrimination; lower perceived risk; and contextual factors such as gender, education, socioeconomic status and family decision-making.^15^ A recent study of predictors of COVID-19 vaccine hesitancy from the UK Household Longitudinal Study found that among vaccine hesitant groups, Black participants were more likely to cite lack of trust in vaccines and worries about unknown future effects of vaccination, whereas Pakistani and Bangladeshi groups were most concerned about side effects as well as unknown future effects.^16^

Studies of COVID-19 vaccination intentions and uptake in HCWs since December 2020 show variability in uptake between countries and, as with general populations, variability by occupational and demographic groups.^8,22,31,32^ A study in a large UK hospital trust showed that 71% of White staff had been vaccinated against COVID-19 as compared to 59% of South Asian staff and 37% of Black staff. Factors associated with vaccine hesitancy (other than belonging to an ethnic minority group) were younger age, female sex and living in more deprived areas.^12^

To date there have been very few large-scale studies of COVID-19 vaccine hesitancy among ethnically diverse HCWs. We undertook an analysis to understand levels of vaccine hesitancy and the factors predicting this in UK HCWs using interim data from the United Kingdom Research study into Ethnicity And COVID-19 outcomes in Healthcare workers (UK-REACH), integrating survey data from a nationwide prospective longitudinal cohort study and qualitative data from HCWs nationwide.

## Methods

### Overview

UK-REACH encompasses six studies to understand the impact of COVID-19 on HCWs from diverse ethnic backgrounds. Here we present data from the baseline questionnaire of the UK-REACH prospective cohort study (administered online from 4^th^ December 2020 with interim data downloaded 19^th^ February 2021), and qualitative data from UK-REACH interviews and focus groups (undertaken from December 2020 to March 2021). Both studies took place in healthcare settings in all four nations of the UK with clinical and non-clinical HCWs from diverse ethnic backgrounds; see study protocols for methodological details.^33,34^

### Prospective nationwide cohort study

Questionnaire design, sampling and baseline questionnaire measures are detailed in the study protocol^33^ and data dictionary (https://www.uk-reach.org/data-dictionary).

#### Study population

All HCWs or ancillary workers in a UK healthcare setting aged 16 or over and/or those registered with one of seven main healthcare regulatory bodies, who responded to an email invitation or who were directly recruited through participating healthcare trusts or open links advertised on social media or in newsletters.

#### Primary outcome measure

We derived the primary outcome, SARS-CoV-2 vaccine hesitancy (binary measure: hesitant versus accepting) from responses to two versions of vaccine questions (VQ1 and VQ2: see Supplementary Figure 1 for details). Vaccine questions were updated during the recruitment/completion period to reflect rapid inception/evolution of the vaccination programme.

#### Predictor variables

We selected variables for inclusion based on the vaccine hesitancy literature, in particular the WHO “Three C’s” model and the vaccine hesitancy determinants matrix,^26^ as well as the UK SAGE report on vaccine hesitancy in ethnic minority groups.^15^ Selected variables measured trust in vaccines and those delivering them; perceived risk of COVID-19; access to vaccines based on job role, sector and location; socio-demographics; and psychological factors. See Supplementary Table 2 for variable list and the data dictionary for details of variables https://www.uk-reach.org/main/data-dictionary/ www.uk-reach.org/main/data-dictionary/. We included a variable to indicate whether participants had answered VQ1 (between 4^th^ and 20^th^ December 2020) or VQ2 (between 21^st^ December 2020 and 19^th^ February 2021). In addition, participants whose VQ2 response indicated that they had considered or were considering not having the vaccine were asked to indicate why they were hesitant.

### Statistical analysis

We summarised categorical variables as count and percentage, and continuous variables as mean (standard deviation [SD]) or median (interquartile range [IQR]) depending on their distribution. We compared groups (hesitant vs accepting, and ethnic groups) with chi-squared tests for categorical variables, and t-tests and analyses of variance for continuous measures, with non-parametric equivalents used as appropriate. Due to the number of tests being performed we considered associations statistically significant at p≤0·001. We checked 2 × 2 interactions with ethnicity on a complete case analysis, which did not significantly improve the model fit.

We used univariable and multivariable logistic regression to determine unadjusted and adjusted associations of variables described above with SARS-CoV-2 vaccine hesitancy.

We used multiple imputation (MI) to replace missing data in all logistic regression models using the package *mice* (Multiple Imputation by Chained Equations) v3·13·0 in *R* version 4·0·4, using predictive mean matching (pmm) for all variables, with 20 imputations and five iterations per imputation (See Supplementary Table 1).

### Qualitative data and analysis

#### Study population

Clinical and non-clinical HCWs aged 16 or older from ethnic minority and White backgrounds with experience of working in UK healthcare settings during COVID-19, recruited through study partners, community organisations, and NHS organisations across the UK.

#### Data analysis

We used framework analysis to analyse anonymised transcripts from interviews and focus groups, and free text data from the cohort study. In interviews and focus groups, we collected data using a piloted topic guide exploring experiences of working during the COVID-19 pandemic, fears and concerns, stigma, discrimination, racism, views on the COVID-19 vaccine, challenges participants encountered in accessing information, and their perceived risk. We collected data on participant gender, ethnicity, age, country of birth, and job role. From the cohort study, free text data were included for participants who discussed SARS-CoV-2 vaccination in response to the following three questions : “What are your thoughts on why people from ethnic minorities working in health and care have been more severely affected by COVID-19?”, “How do you see society changing as a result of COVID-19?” and “How do you see your own future changing as a result of COVID-19?”^35^

We developed the initial framework based on a preliminary analysis of the data and the WHO framework for behavioural considerations for acceptance and uptake of COVID-19 vaccines.^36^ We piloted the framework with the first five transcripts, and refined it iteratively during analysis. The framework encompasses “Drivers of vaccine hesitancy” relating to “health information and messaging”, “Motivation” utilising the ‘Three Cs Model’, and “Improving delivery”.

### Ethical approval

Both studies were approved by the Health Research Authority (Brighton and Sussex Research Ethics Committee; ethics reference: 20/HRA/4718). All participants gave written informed consent.

### Involvement and engagement

We worked closely with a Professional Expert Panel of HCWs from a range of ethnic backgrounds, occupations, and genders, as well as with national and local organisations (see study protocols).^33,34^

## Results

### Prospective nationwide cohort study

#### Description of analysed cohort

Between 4^th^ December 2020 and 19^th^ February 2021, professional regulators sent 1,052,875 HCWs email invitations with a link to the questionnaire, and 21 National Health Service (NHS) Hospital Trusts publicised the questionnaire to their staff and invited staff by email. As of 19^th^ February 2021, 15,151 participants had started the questionnaire. The analysed interim cohort were formed of 11,584 participants who both completed the questionnaire and answered the question about their sex. See Table 1 for the cohort demographics, job type and location.

#### Univariable results

Table 1 shows the cohort stratified by SARS-CoV-2 vaccine hesitancy; Table 2 shows univariable relationships between hesitancy and vaccine-related trust and perceived risk of COVID-19. Briefly, just under a quarter of participants (2694/11,584; 23·3%) were vaccine hesitant. Over half (51·0%) of Black Caribbean, 38·1% of Mixed White and Black Caribbean, 34·4% of Black African, 32·4% of Chinese, 29·8% of Pakistani, and 28·7% of the White Other^1^ group were vaccine hesitant, compared to 21·0% of White British, 19·6% of Indian and 18·8% of Bangladeshi HCWs. The least hesitant occupational group was the Doctors and medical support group (18·4% hesitant) and the most hesitant was the Nursing, Nursing associates and Midwives group (28·2% hesitant).

**Table 1:** Demographic characteristics of cohort stratified by SARS-CoV-2 vaccine hesitancy.

| Variable | Total<br>(n=11,584) | SARS-CoV-2 vaccine |  |  |
| --- | --- | --- | --- | --- |
|  |  | Not hesitant<br>8691 (75.0%) | Hesitant<br>2694 (23.3%) | Missing<br>199 (1.7%) |
| **Age, median (IQR)** | 45 (34 - 54) | 46 (36 - 55) | 40 (31 - 51) | 43 (34 - 54) |
| **Sex, n(%)** |  |  |  |  |
| Male | 2797 (24.2%) | 2298 (26.4%) | 466 (17.3%) | 33 (16.6%) |
| Female | 8787 (75.9%) | 6393 (73.6%) | 2228 (82.7%) | 166 (83.4%) |
| **Ethnicity, n(%)** |  |  |  |  |
| White - English, Welsh, Scottish, Northern Irish | 6907 (60.8%) | 5365 (62.8%) | 1452 (55.2%) | 90 (46.9%) |
| White - Irish | 209 (1.8%) | 156 (1.8%) | 50 (1.9%) | <5 (<2%) |
| White - Other/Gypsy Irish Traveller | 878 (7.7%) | 606 (7.1%) | 252 (9.6%) | 20 (10.4%) |
| Asian - Indian | 1187 (10.4%) | 936 (11.0%) | 232 (8.8%) | 19 (9.9%) |
| Asian - Pakistani | 315 (2.8%) | 215 (2.5%) | 94 (3.6%) | 6 (3.1%) |
| Asian - Bangladeshi | 69 (0.6%) | 55 (0.6%) | 13 (0.5%) | <5 (<2%) |
| Asian - Chinese | 253 (2.2%) | 166 (1.9%) | 82 (3.1%) | 5 (2.6%) |
| Asian - Other | 365 (3.2%) | 269 (3.2%) | 89 (3.4%) | 7 (3.7%) |
| Black - African | 349 (3.1%) | 210 (2.5%) | 120 (4.6%) | 19 (9.9%) |
| Black - Caribbean | 102 (0.9%) | 44 (0.5%) | 52 (2.0%) | 6 (3.1%) |
| Black - Other | 20 (0.2%) | 11 (0.1%) | 7 (0.3%) | <5 (<2%) |
| Mixed - White & Black African | 66 (0.6%) | 47 (0.6%) | 18 (0.7%) | <5 (<2%) |
| Mixed - White & Black Caribbean | 84 (0.7%) | 50 (0.6%) | 32 (1.2%) | <5 (<2%) |
| Mixed - White & Asian | 179 (1.6%) | 140 (1.6%) | 37 (1.4%) | <5 (<2%) |
| Mixed - Other | 142 (1.3%) | 103 (1.2%) | 36 (1.4%) | <5 (<2%) |
| Other - Arab | 122 (1.1%) | 83 (1.0%) | 34 (1.3%) | 5 (2.6%) |
| Other | 123 (1.1%) | 89 (1.0%) | 33 (1.3%) | <5 (<2%) |
| **Religion, n(%)** |  |  |  |  |
| None | 3939 (36.0%) | 2995 (36.4%) | 896 (35.6%) | 48 (25.8%) |
| Christian | 5109 (46.7%) | 3827 (46.5%) | 1176 (46.7%) | 106 (57.0%) |
| Buddhist | 133 (1.2%) | 93 (1.1%) | 36 (1.4%) | <5 (<2%) |
| Hindu | 697 (6.4%) | 557 (6.8%) | 132 (5.2%) | 8 (4.3%) |
| Jewish | 107 (1.0%) | 93 (1.1%) | 13 (0.5%) | <5 (<2%) |
| Muslim | 670 (6.1%) | 472 (5.7%) | 184 (7.3%) | 14 (7.5%) |
| Sikh | 120 (1.1%) | 89 (1.1%) | 28 (1.1%) | <5 (<2%) |
| Other | 156 (1.4%) | 99 (1.2%) | 55 (2.2%) | <5 (<2%) |
| **Country of birth, n(%)** |  |  |  |  |
| UK | 8335 (73.4%) | 6411 (73.9%) | 1924 (71.7%) | 127 (63.8%) |
| Outside UK | 3024 (26.6%) | 2264 (26.1%) | 760 (28.3%) | 72 (36.2%) |
| **Job role, n(%)** |  |  |  |  |
| Doctors and medical support | 2679 (24.0%) | 2166 (25.9%) | 493 (19.1%) | 20 (10.7%) |
| Nurses, NAs, Midwives | 2300 (20.6%) | 1598 (19.1%) | 648 (25.1%) | 54 (28.9%) |
| Allied Health Professionals | 4959 (44.5%) | 3689 (44.1%) | 1184 (45.9%) | 86 (46.0%) |
| Dental | 716 (6.4%) | 551 (6.6%) | 151 (5.9%) | 14 (7.5%) |
| Administrative/Estates/Other | 488 (4.4%) | 369 (4.4%) | 106 (4.1%) | 13 (7.0%) |
| **Job location, n(%)** |  |  |  |  |
| Not in hospital | 5013 (44.5%) | 3859 (45.7%) | 1072 (40.8%) | 82 (44.1%) |
| Hospital | 6254 (55.5%) | 4595 (54.4%) | 1555 (59.2%) | 104 (55.9%) |
| **IMD quintile** |  |  |  |  |
| 1 (most deprived) | 965 (9.5%) | 661 (8.7%) | 281 (12.0%) | 23 (12.8%) |
| 2 | 1660 (16.4%) | 1169 (15.4%) | 458 (19.5%) | 33 (18.3%) |
| 3 | 2097 (20.7%) | 1574 (20.7%) | 479 (20.4%) | 44 (24.4%) |
| 4 | 2478 (24.5%) | 1900 (25.0%) | 527 (22.4%) | 51 (28.3%) |
| 5 (least deprived) | 2926 (28.9%) | 2294 (30.2%) | 603 (25.7%) | 29 (16.1%) |

**Table 2:** Selected predictor variables stratified by SARS-CoV-2 vaccine hesitancy. For categorical variables, percentages are column wise apart from totals which are computed row wise. For details of the derivation of the trust variables please see supplementary information. Comorbidities Include: organ transplant, diabetes, heart disease, hypertension, stroke, kidney disease, liver disease, anaemia, asthma, lung disease, cancer, neurological disorder and immunosuppression.

| Variable | Total<br>(n=11,584) | SARS-CoV-2 vaccine |  |  | P value |
| --- | --- | --- | --- | --- | --- |
|  |  | Not hesitant<br>8691 (75.0%) | Hesitant<br>2696 (23.3%) | Missing<br>199 (1.7%) |  |
| TRUST VARIABLES |  |  |  |  |  |
| Belief in COVID-19 ‘conspiracies’ score, med (IQR)<br>(min 6 [does not believe] - max 24 [strongly believes]) | 9 (8 - 10) | 8 (7 - 10) | 10 (8 - 11) | 10 (9 - 13) | <0.0001* |
| Pro-vaccine score, med (IQR)<br>(min 4 [anti-vaccination] - max 20 [pro-vaccination]) | 16 (14 - 17) | 16 (14 - 17) | 14 (12 - 16) | 13 (11 - 16) | <0.0001* |
| Influenza vaccination status 2019 - 2020, n(%)<br>Vaccinated<br>Unvaccinated | 8279 (71.9%)<br>3233 (28.1%) | 6569 (76.0%)<br>2070 (24.0%) | 1605 (59.9%)<br>1074 (40.1%) | 105 (54.1%)<br>89 (45.9%) | <0.0001* |
| Trust in employer to deal with a concern about unsafe clinical practice, n(%)<br>1 (does not trust employer)<br>2<br>3<br>4<br>5 (trusts employer) | 356 (3.3%)<br>950 (8.9%)<br>1807 (16.9%)<br>4113 (38.4%)<br>3481 (32.5%) | 252 (3.1%)<br>649 (8.1%)<br>1241 (15.4%)<br>3113 (38.7%)<br>2781 (34.6%) | 95 (3.8%)<br>289 (11.6%)<br>532 (21.3%)<br>942 (37.8%)<br>636 (25.5%) | 9 (5.1%)<br>12 (6.8%)<br>34 (19.2%)<br>58 (32.8%)<br>64 (36.2%) | <0.0001<sup>†</sup> |
| Discrimination at work on the basis of ethnicity, nationality or religion, n(%)<br>Has not experienced discrimination<br>Has experienced discrimination | 9270 (86.6%)<br>1434 (13.4%) | 7072 (87.9%)<br>977 (12.1%) | 2063 (83.1%)<br>420 (16.9%) | 135 (78.5%)<br>37 (21.5%) | <0.0001* |
| RISK VARIABLES |  |  |  |  |  |
| Previous laboratory evidence of COVID-19 (PCR or serology), n(%)<br>Never tested<br>Tested negative<br>Tested positive | 1903 (16.5%)<br>7350 (63.5%)<br>2316 (20.0%) | 1449 (16.6%)<br>5635 (64.9%)<br>1597 (18.4%) | 420 (15.6%)<br>1610 (59.8%)<br>662 (24.6%) | 34 (17.4%)<br>105 (53.6%)<br>57 (29.1%) | <0.0001* |
| Number of comorbidities, n(%)<br>0<br>1<br>≥2 | 7841 (70.7%)<br>2528 (22.8%)<br>724 (6.5%) | 5801 (69.6%)<br>1956 (23.5%)<br>580 (7.0%) | 1910 (74.1%)<br>532 (20.6%)<br>137 (5.3%) | 130 (73.5%)<br>40 (22.6%)<br>7 (4.0%) | <0.0001* |
| Pregnancy, n(%)<br>Not pregnant<br>Pregnant | 10,948 (98.7%)<br>141 (1.3%) | 8286 (99.4%)<br>50 (0.6%) | 2490 (96.7%)<br>86 (3.3%) | 194 (97.5%)<br>5 (2.5%) | <0.0001* |
| Perceived risk of hospitalisation with COVID-19, med, (IQR)<br>(100 point scale) | 20 (5 - 50) | 20 (5 - 50) | 20 (5 - 40) | 15 (3 - 50) | 0.0004<sup>†</sup> |
| Concerned about unknowingly spreading COVID-19, n(%)<br>Not concerned<br>Concerned | 5745 (49.8%)<br>5781 (50.2%) | 4285 (49.5%)<br>4367 (50.5%) | 1362 (50.8%)<br>1319 (49.2%) | 98 (50.8%)<br>95 (49.2%) | 0.2* |
| Exposed to COVID-19 patients at work, n(%)<br>Unexposed<br>Exposed | 7164 (66.1%)<br>3682 (34.0%) | 5476 (67.3%)<br>2663 (32.7%) | 1589 (62.9%)<br>939 (37.1%) | 99 (55.3%)<br>80 (44.7%) | <0.0001* |

\*chi-square, <sup>†</sup>Wilcoxon rank-sum for comparison between hesitant and non-hesitant cohorts.

### Error! Reference source not found

Hesitant participants scored higher on the’COVID-19 conspiracy beliefs’ scale (median hesitant: 10, IQR: 8-11; non-hesitant: 8, 7-10 p<0·0001), were less confident their employer would address a concern about unsafe clinical practice (63·3% vs 73·3% p<0·001) and were more likely to have laboratory evidence of previous SARS-CoV-2 infection (24·6% vs 18·4%, p<0·001) compared to the non-hesitant cohort. 136 pregnant HCWs were included in the analysis, of whom 86 (63·2%) were SARS-CoV-2 vaccine hesitant. Supplementary Table 4 shows vaccine-related trust and risk factors stratified by ethnicity.

#### Reasons for hesitancy

Among those who reported reasons for SARS-CoV-2 vaccine hesitancy, the White group (White British, White Irish, White Other and White Gypsy/Irish Traveller) were less concerned than other ethnic groups about potential vaccine side effects or about the vaccine not having been tested in diverse ethnic groups, and they were less likely to want to delay until others had the vaccine. Reasons for hesitancy overall and by broad ethnic grouping are given in Supplementary Table 3.

**Table 3:** Unadjusted and adjusted analysis of SARS-CoV-2 vaccine hesitancy predictors.

| Variable | OR (95% CI) | p value | aOR (95% CI) | p value |
| --- | --- | --- | --- | --- |
| **Age** (for each decade increase) | 0.71 (0.69 - 0.74) | <0.001 | 0.74 (0.70 - 0.78) | <0.001 |
| **Sex** |  |  |  |  |
| Male | Ref | - | Ref | - |
| Female | 1.72 (1.54 - 1.93) | <0.001 | 1.42 (1.24 - 1.62) | <0.001 |
| **Ethnicity** |  |  |  |  |
| White - British | Ref | - | Ref | - |
| White - Irish | 1.18 (0.85 - 1.63) | 0.32 | 1.39 (0.96 - 2.02) | 0.08 |
| White - Other/Gypsy Irish Traveller | 1.55 (1.33 - 1.81) | <0.001 | 1.48 (1.19 - 1.84) | 0.001 |
| Asian - Indian | 0.92 (0.79 - 1.07) | 0.28 | 0.76 (0.57 - 1.02) | 0.07 |
| Asian - Pakistani | 1.62 (1.26 - 2.08) | <0.001 | 1.18 (0.78 - 1.79) | 0.42 |
| Asian - Bangladeshi | 0.87 (0.47 - 1.59) | 0.64 | 0.66 (0.32 - 1.39) | 0.28 |
| Asian - Chinese | 1.80 (1.37 - 2.36) | <0.001 | 1.59 (1.15 - 2.20) | 0.005 |
| Asian - Other | 1.23 (0.96 - 1.57) | 0.1 | 1.03 (0.74 - 1.42) | 0.86 |
| Black - African | 2.09 (1.66 - 2.63) | <0.001 | 2.05 (1.49 - 2.82) | <0.001 |
| Black - Caribbean | 3.91 (2.62 - 5.84) | <0.001 | 3.37 (2.11 - 5.37) | <0.001 |
| Black - Other | 2.45 (0.99 - 6.06) | 0.05 | 1.63 (0.52 - 5.06) | 0.40 |
| Mixed - White & Black Caribbean | 2.23 (1.43 - 3.48) | <0.001 | 1.62 (0.98 - 2.67) | 0.06 |
| Mixed - White & Black African | 1.35 (0.78 - 2.33) | 0.28 | 1.36 (0.87 - 2.11) | 0.33 |
| Mixed - White & Asian | 0.95 (0.66 - 1.38) | 0.79 | 0.89 (0.59 - 1.36) | 0.60 |
| Mixed - Other | 1.29 (0.88 - 1.90) | 0.19 | 1.35 (0.87 - 2.11) | 0.18 |
| Arab | 1.43 (0.96 - 2.13) | 0.08 | 1.65 (0.97 - 2.82) | 0.07 |
| Other | 1.36 (0.91 - 2.03) | 0.13 | 1.41 (0.88 - 2.26) | 0.15 |
| **Job role** |  |  |  |  |
| Doctors and medical support | Ref | - | Ref | - |
| Nurses, NAs, Midwives | 1.75 (1.54 - 2.00) | <0.001 | 1.17 (0.98 - 1.41) | 0.08 |
| Allied Health Professionals | 1.39 (1.24 - 1.57) | <0.001 | 0.99 (0.85 - 1.16) | 0.90 |
| Dental | 1.21 (0.99 - 1.48) | 0.06 | 0.75 (0.58 - 0.97) | 0.03 |
| Admin / estates / other | 1.25 (0.99 - 1.57) | 0.06 | 1.03 (0.78 - 1.36) | 0.86 |
| **Job location** |  |  |  |  |
| Not in hospital | Ref | - | Ref | - |
| Hospital | 1.22 (1.12 - 1.34) | <0.001 | 1.18 (1.06 - 1.32) | 0.004 |
| **Religion** |  |  |  |  |
| No religion | Ref | - | Ref | - |
| Christian | 1.03 (0.94 - 1.14) | 0.52 | 0.99 (0.87 - 1.12) | 0.85 |
| Buddhist | 1.27 (0.86 - 1.86) | 0.23 | 1.10 (0.70 - 1.72) | 0.68 |
| Hindu | 0.83 (0.68 - 1.02) | 0.08 | 1.10 (0.79 - 1.53) | 0.58 |
| Jewish | 0.49 (0.27 - 0.88) | 0.02 | 0.54 (0.28 - 1.03) | 0.06 |
| Muslim | 1.31 (1.09 - 1.58) | 0.003 | 1.02 (0.73 - 1.42) | 0.92 |
| Sikh | 1.10 (0.72 - 1.68) | 0.67 | 1.39 (0.81 - 2.38) | 0.24 |
| Other | 1.74 (1.25 - 2.44) | 0.001 | 1.77 (1.19 - 2.62) | 0.005 |
| **Religiosity** | 1.10 (1.05 - 1.15) | <0.001 | 1.03 (0.97 - 1.10) | 0.38 |
| **Country of Birth** |  |  |  |  |
| Outside UK | Ref | - | Ref | - |
| UK | 0.89 (0.80 - 0.97) | 0.01 | 1.26 (1.07 - 1.48) | 0.006 |
| **IMD quintile** |  |  |  |  |
| 1 (most deprived) | 1.38 (1.16 - 1.64) | <0.001 | 0.96 (0.79 - 1.17) | 0.66 |
| 2 | 1.26 (1.08 - 1.46) | 0.003 | 1.10 (0.93 - 1.31) | 0.26 |
| 3 | Ref | - | Ref | - |
| 4 | 0.93 (0.81 - 1.07) | 0.32 | 1.01 (0.86 - 1.18) | 0.90 |
| 5 (least deprived) | 0.87 (0.76 - 0.99) | 0.04 | 1.00 (0.87 - 1.16) | 0.95 |
| **Influenza vaccination 2019 - 2020** |  |  |  |  |
| Unvaccinated | Ref | - | Ref | - |
| Vaccinated | 0.46 (0.43 - 0.51) | <0.001 | 0.51 (0.46 - 0.57) | <0.001 |
| **Pro-vaccine attitudes** | 0.78 (0.73 - 0.84) | <0.001 | 0.82 (0.78 - 0.86) | <0.001 |
| **COVID-19 conspiracy beliefs scale** | 1.22 (1.20 - 1.24) | <0.001 | 1.12 (1.08 - 1.16) | <0.001 |
| **Trust in employer:**<br>secure raising concerns<br>confident concerns would be addressed | 0.85 (0.81 - 0.88)<br>0.82 (0.78 - 0.85) | <0.001<br><0.001 | 1.02 (0.96 - 1.09)<br>0.87 (0.82 - 0.93) | 0.51<br><0.001 |
| **Discrimination at work** on the basis of ethnicity, nationality or religion | 1.45 (1.28 - 1.64) | <0.001 | 0.99 (0.84 - 1.17) | 0.93 |
| **Number of comorbidities**<br>0<br>1<br>≥2 | Ref<br>0.83 (0.75 - 0.93)<br>0.72 (0.59 - 0.87) | -<br>0.001<br>0.001 | Ref<br>0.97 (0.85 - 1.10)<br>1.11 (0.87 - 1.40) | -<br>0.65<br>0.40 |
| **BMI category**<br><18.5<br>18.5 to <25<br>25 to <30<br>30 to <40<br>≥ 40 | 1.04 (0.72 - 1.52)<br>Ref<br>0.91 (0.82 - 1.01)<br>0.96 (0.85 - 1.09)<br>0.87 (0.65 - 1.16) | 0.81<br>-<br>0.07<br>0.52<br>0.33 | 0.89 (0.58 - 1.36)<br>Ref<br>0.90 (0.80 - 1.02)<br>0.87 (0.76 - 1.01)<br>0.68 (0.50 - 0.95) | 0.58<br>-<br>0.09<br>0.07<br>0.02 |
| **Pregnancy**<br>Not pregnant<br>Pregnant | Ref<br>5.87 (4.14 - 8.33) | -<br><0.001 | Ref<br>7.12 (4.74 - 10.70) | -<br><0.001 |
| **Previous evidence of COVID-19 (PCR or serology)**<br>Negative<br>Never tested<br>Positive | Ref<br>1.02 (0.90 - 1.15)<br>1.46 (1.32 - 1.62) | -<br>0.76<br><0.001 | Ref<br>0.95 (0.83 - 1.10)<br>1.30 (1.14 - 1.47) | -<br>0.52<br><0.001 |
| **Perceived risk of hospitalisation with COVID-19** (for each 10 point increase) | 0.97 (0.95 - 0.99) | 0.001 | 0.97 (0.94 - 0.99) | 0.009 |
| **Perceived risk of unknowingly spreading COVID-19**<br>Not concerned<br>Quite or very concerned | Ref<br>0.95 (0.87 - 1.03) | -<br>0.22 | Ref<br>0.88 (0.79 - 0.97) | -<br>0.01 |
| **Exposure to COVID-19 patients at work**<br>Unexposed<br>Exposed | Ref<br>1.22 (1.11 - 1.34) | -<br><0.001 | Ref<br>0.90 (0.80 - 1.01) | -<br>0.07 |
| **Personality factors**<br>Agreeableness<br>Conscientiousness<br>Extraversion<br>Neuroticism<br>Openness | 0.97 (0.96 - 0.99)<br>0.99 (0.98 - 1.01)<br>0.98 (0.97 - 0.99)<br>1.02 (1.01 - 1.04)<br>0.99 (0.97 - 1.00) | 0.001<br>0.40<br>0.003<br><0.001<br>0.03 | 0.98 (0.96 - 0.99)<br>1.00 (0.98 - 1.02)<br>0.99 (0.98 - 1.01)<br>0.99 (0.98 - 1.00)<br>0.99 (0.98 - 1.01) | 0.01<br>0.75<br>0.04<br>0.44<br>0.23 |
| **Fatalism** | 1.03 (1.02 - 1.04) | <0.001 | 1.00 (0.99 - 1.01) | 0.54 |
| **Information sources**<br>Friends<br>Mainstream media<br>Official<br>Scientific | 1.40 (1.19 - 1.65)<br>0.68 (0.60 - 0.76)<br>0.77 (0.63 - 0.94)<br>0.88 (0.76 - 1.02) | <0.001<br><0.001<br>0.01<br>0.09 | 1.27 (1.02 - 1.57)<br>0.82 (0.70 - 0.96)<br>0.85 (0.66 - 1.10)<br>1.28 (1.07 - 1.53) | 0.03<br>0.01<br>0.21<br>0.007 |
| **Time of questionnaire completion**<br>December 2020 or before<br>January 2021 or after | Ref<br>0.49 (0.45 - 0.54) | - | Ref<br>0.52 (0.30 - 0.90) | -<br><0.001 |

##### Multivariable results

##### Demographic predictors of SARS-CoV-2 vaccine hesitancy

Table 3 shows univariable and multivariable logistic regression models with an outcome of SARS-CoV-2 vaccine hesitancy. After adjusting for socio-demographic, job, trust, perceived COVID-19 risk, and psychological factors, vaccine hesitancy was less likely with increasing age (aOR 0·74 95%CI 0·70–0·78 for each decade increase) and more likely among female HCWs (aOR 1·42 95%CI 1·24–1·62). Compared to White British HCWs, those from Black Caribbean (aOR 3·37 95%CI 2·11– 5·37), Black African (aOR 2·05, 95%CI 1·49–2·82), and White Other (aOR 1·48 95%CI 1·19–1·84) ethnic groups were significantly more likely to be vaccine hesitant.

##### Trust, COVID-19 risk and psychological predictors of SARS-CoV-2 vaccine hesitancy and refusal

Greater belief in COVID-19 conspiracies was significantly associated with increased odds of hesitancy (aOR 1·12, 95%CI 1·08–1·16 for each 1 point increase on the scale). Increasing confidence that concerns raised about unsafe practice would be addressed by their employer decreased the odds of hesitancy (aOR 0·87, 95%CI 0·82 - 0·93). Those who had received the influenza vaccine in winter 2019/2020 were around half as likely to be SARS-CoV-2 vaccine hesitant compared to those who had not (aOR 0·51, 95%CI 0·46 - 0·57). HCWs who reported testing positive for SARS-CoV-2 by PCR or serology, were significantly more likely to be hesitant than those testing negative (aOR 1·30, 95%CI 1·14 - 1·47). Pregnant HCWs were over 7 times as likely to be hesitant (aOR 7·12, 95%CI 4·74 - 10·70).

### Qualitative study

#### Description of sample

We included 99 individuals, 41 recruited through interviews (n=24) and focus groups (n=17), and 58 from the longitudinal cohort study (free text comments provided about vaccinations). Among the 41 qualitative participants, 13 were Asian (32%), 12 were Black (29%), and 10 were White (24%). 27 (66%) were women, and 24 were born in the UK (59%). 18 participants were allied health professionals, pharmacists, and dentists (44%), whilst 9 were doctors (22%), 3 were nurses or midwives (7%), and 11 were non-clinical (27%). Among the 58 cohort participants, 42 were White (72%), 8 were Asian (14%), and 4 were Black (7%). 48 participants (83%) were women, and 44 (76%) were born in the UK. 26 (45%) of participants were allied health professionals, pharmacists, or dentists, 7 were doctors (12%), whilst 23 (40%) were nurses or midwives.

#### Drivers of vaccine hesitancy

We identified four intersecting themes describing key drivers of and ways to address vaccine hesitancy among HCWs: Trust, Perceived risk, Health information and messaging, and Improving delivery (See Figure 1; Supplementary Tables 5-8 for quotes).

**Figure 1:**
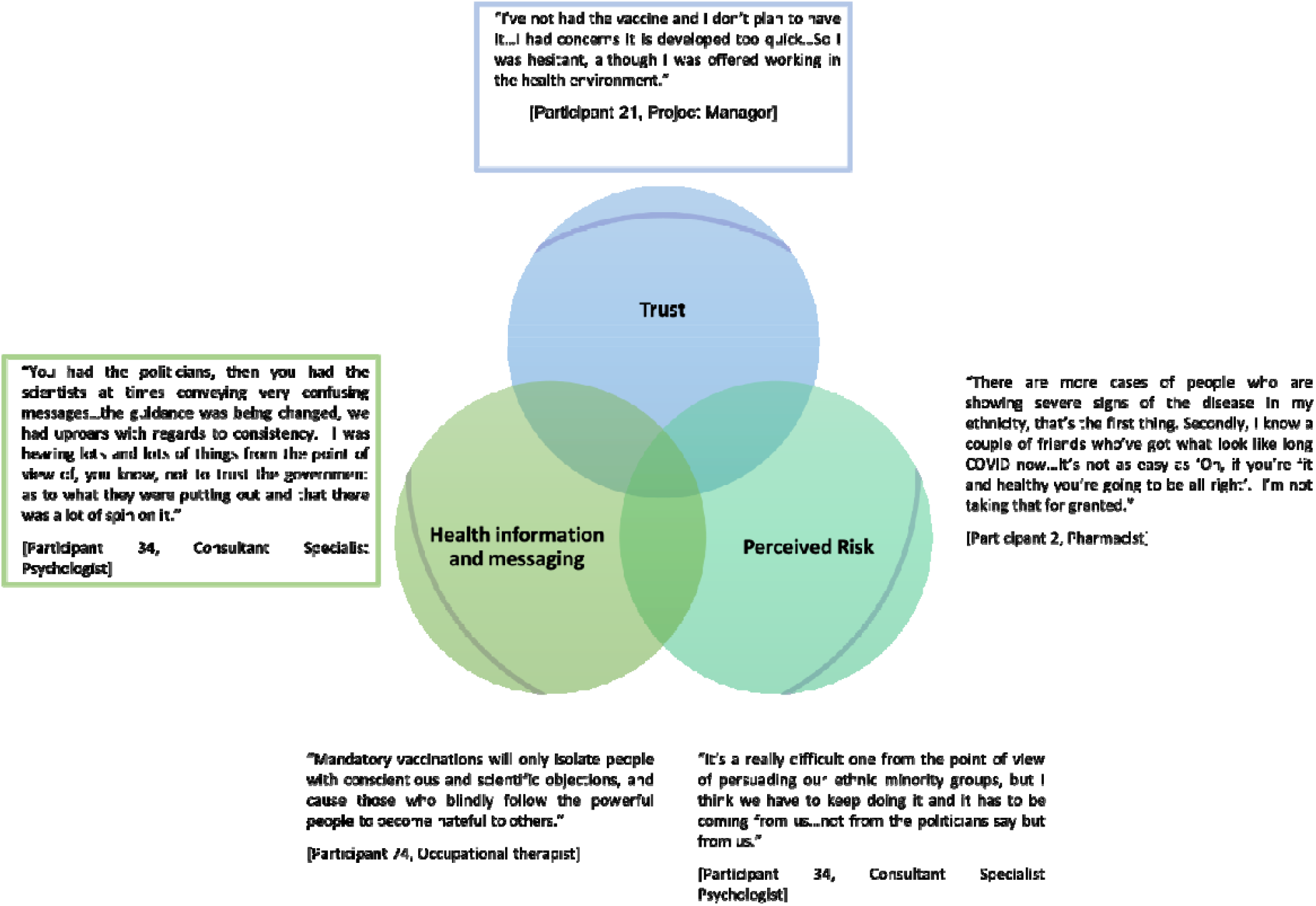
Qualitative themes.

##### Trust

Participants described their enthusiasm about the vaccine, appreciation of being prioritised, and the role of trust in colleagues, the NHS, and health information in facilitating vaccine uptake. Narratives also highlighted the influence of experiences of discrimination and structural inequities on trust and vaccine hesitancy, and the ubiquity of concerns around the vaccine across both those who declined to be vaccinated and those who described themselves as pro-vaccine.

##### Trust in vaccinations

Whilst some participants described a lack of confidence in vaccines generally, most participants described being accepting of routine or flu vaccinations. Key concerns for the COVID-19 vaccine related to speed of development, lack of longitudinal data, and potential side effects, as well as efficacy against SARS-CoV-2 variants. There were also concerns about the underrepresentation of individuals from ethnic minority backgrounds in vaccine trials.

##### Trust in those producing, giving, and taking vaccines

Vaccine confidence among colleagues, family, friends and community members increased HCW trust in the vaccine. More senior colleagues - particularly clinicians - were especially influential, and conversely trust was eroded when they did not adhere to guidance. Some participants described a contradiction between their own concerns around having the vaccine, yet promoting it for the wider public through their roles.

##### Perceived risks of COVID-19 to self and others

Whilst some participants felt at low risk, others expressed concern about the risk of exposure in their role and fears of having COVID-19, even if they did not have other key risk factors. Previous infection, knowing people who had been unwell or passed away from COVID-19, and concerns about infection of their families and loved ones often increased perceived risk. Participants’ views about the extent to which vaccination could reduce risk also influenced their decision to be vaccinated, as did their desire to reduce their risk of transmission and protect their close contacts. Participants also discussed how they perceived ethnicity to influence risk. Whilst prioritisation of NHS workers for vaccines was welcomed, some felt ethnic minority groups should have been prioritised given their increased risk.

##### Health information and messaging

Accessibility and trustworthiness of health information shaped vaccine concerns. Complex information, conflicting and changing guidance, overwhelming amounts of material, and poor provision of information in other languages contributed to a lack of trust, confusion, and ultimately vaccine hesitancy. Participants also noted the stigma around vaccine hesitancy and lack of vaccine knowledge.

Participants obtained information from numerous sources. Social media was often described as potentially misleading or unreliable, but some participants acknowledged its usefulness for raising awareness, vaccine promotion, and disseminating messaging, especially as information shared by community members may be more trusted. Participants also frequently accessed information through the news, or Government and NHS sources. However, the positive presentation of vaccines by these sources was felt by some to be insincere with potential risks not being transparently communicated. This fed into suspicions around official reports on COVID-19 further contributing to HCW mistrust.

There were varying responses to the focus on ethnic minorities. While prioritisation of NHS workers for vaccines was welcomed, some felt HCWs from ethnic minority backgrounds should have been further prioritised given evidence of the disproportionate impact of COVID-19 on these communities. The narratives also illustrated discomfort with the focus on ethnic minorities in the media, messaging and vaccine promotion campaigns, which singled out these communities as’vaccine hesitant’ and increased stigma. One participant brought attention to discourse around reported low vaccine uptake of the vaccine among Black doctors, calling for greater transparency and accuracy around uptake rates, and better understandings of the factors that inform decisions about vaccines.

##### Inclusive communication

Participants highlighted the value of communicating messages through a range of media and languages, and engaging directly with people to respond to questions or concerns, and tackle misinformation. Participants also advocated for using existing resources such as multilinguistic healthcare staff to strengthen the accessibility and trustworthiness of health information.

Participants also described the importance of language in how groups are described, and the need to avoid assumptions or stereotyping associated with ethnicity. This was important for creating more inclusive communication around how at-risk groups - and ethnic minority communities in particular - are described in research, the workplace, and the media.

##### Increasing transparency and trust

Trust and informed decision-making about vaccines was influenced by how risk groups were identified and prioritised, who was eligible, and the perceived risks and benefits. Participants explained the importance of transparent and clear communication through hospital Trusts.

##### Equity, opportunity and mandatory vaccination

Participants underscored the need to ensure equity in vaccine delivery, with some advocating prioritisation of staff experiencing the greatest barriers to getting the vaccine, or who were at greatest risk. Whilst some participants advocated *for “mandatory vaccinations for those choosing to work in health and social care settings”* (Participant 84, Speech and language therapist), others were concerned about the potential lack of equity for those who chose not to have the vaccine, and that mandating vaccination could create further ethnic divides between communities and increase stigma and discrimination. Participants also discussed how ensuring equity in accessibility and opportunity to have the vaccine would be paramount for improving delivery.

##### Outreach through involvement

Participants described how the vaccine roll-out could be improved through better engagement with and involvement of HCWs, particularly those from ethnic minority communities. The narratives pointed to the lack of inclusion of marginalised communities throughout the pandemic, and the potential benefit of increasing visibility of less well represented groups in the media to promote vaccine uptake and trust.

Participants also discussed the importance of promoting vaccination through trusted networks, and the value of more proactive involvement and engagement of healthcare workers from diverse ethnic backgrounds. An important aspect of both building trust and increasing accessibility was acknowledging cultural differences in understandings of and access to vaccines. Participants highlighted how the involvement of minoritised communities can play an important role in bridging cultural divides, and the potential benefit of outreach activities for addressing logistical challenges in delivering the vaccine.

## Discussion

In this analysis of interim data from nearly 12,000 HCWs across the UK, approximately a quarter of participants reported SARS-CoV-2 vaccine hesitancy. HCWs from Black Caribbean, Black African and White Other ethnic groups reported higher hesitancy than those from the White British group after adjusting for other predictors. Additional factors predicting hesitancy were scoring higher on the COVID-19 conspiracy beliefs scale, lower trust in employer, pregnancy, and previous COVID-19 infection. Qualitative data showed information and messaging influenced vaccine concerns. Speed of vaccine development, experiences of discrimination and structural inequalities also contributed to a lack of trust in the vaccine.

Our finding of 23% of UK HCWs being SARS-CoV-2 vaccine hesitant is in keeping with a recent systematic review of COVID-19 vaccine uptake, which found an average acceptance of 57% (range 28%-78%) across countries and occupation groups.^8^ Many smaller SARS-CoV-2 vaccine hesitancy studies have been conducted outside the UK with common predictors of vaccine hesitancy being female sex, non-medical occupation, lack of influenza vaccination and lower perceived risk of COVID-19.^6,7,37–39^ There is conflicting evidence regarding the effect of age and previous COVID-19 on SARS-CoV-2 vaccine hesitancy.^37,40–42^ Importantly, only two studies, both conducted in the US, examined the impact of ethnicity on SARS-CoV-2 vaccine hesitancy after adjustment for confounders, with both finding that Black ethnic groups were more likely to be hesitant compared to White HCWs.^41,42^ Whilst data on vaccine hesitancy in UK HCWs are limited, recent work examining vaccine uptake amongst hospital staff in the UK found that 35·5% of HCWs had not been vaccinated; vaccination rates were highest amongst White HCWs and, in-line with our findings, lowest among Black ethnic groups.^12^

Due to the novel nature of COVID-19, the evidence base for barriers to SARS-CoV-2 vaccination in ethnic minority communities is limited. However, UK’s SAGE ethnicity subgroup identified barriers to vaccine uptake amongst ethnic minority groups including lower trust in vaccine efficacy/safety (particularly speed of vaccine development), mistrust of healthcare organisations (due to prior unethical research practices), lack of representation in vaccine trials, and institutional racism and discrimination.^30^ Our study provides evidence that these same factors may influence vaccine hesitancy in HCWs. Many of these themes emerged in our qualitative data, with HCWs describing reservations about accepting vaccination rooted in safety concerns due to the short development timeframe of current SARS-CoV-2 vaccines. Experiences of health inequities and knowledge of historic unethical health and research practices were cited by some Black HCWs as influencing their mistrust of the NHS. This overarching mistrust in the organisation was also reflected in attitudes towards SARS-CoV-2 vaccination with a perception of low ethnic minority involvement in trials to gauge vaccine safety/efficacy, and the lack of prioritisation within the vaccination rollout despite evidence of the disproportionate impact on the health of those from minority ethnic backgrounds. Additionally, in data from the cohort study, lower trust in employer was found to predict hesitancy, and high proportions of vaccine hesitant ethnic minority HCWs expressed concerns regarding vaccine safety and about a lack of testing in all ethnic groups.

These results have important implications for public health measures aimed at improving vaccine uptake. It has been reported that mandatory SARS-CoV-2 vaccination is being considered for care home staff in the UK,^43^ and the Italian government has mandated vaccination in HCWs (with those that refuse offered duties that do not risk viral transmission or suspension without pay).^44^ Whilst these measures may improve vaccine uptake, our results indicate that implementing these policies may undermine trust (both in the employing healthcare organisations and in the vaccination programme).^45^ Given that this effect would not be seen equally across ethnic groups, such interventions have the potential to increase stigma and discrimination and widen ethnic disparities.

We found that higher scores on the COVID-19 conspiracy beliefs scale was associated with vaccine hesitancy, and this was also more likely in ethnic minority groups as compared to those of White ethnicity. To our knowledge, we are the first to show this effect in a HCW population. A general population survey in the UK found that belief in COVID-19 conspiracies was more likely in those who were SARS-CoV-2 vaccine hesitant and in ethnic minority groups.^46^ Our findings confirm that misinformation relating to COVID-19 is important even amongst HCWs, and strategies to tackle this may increase vaccination uptake amongst HCWs and the population at large.

We found that those with evidence of previous COVID-19 were more likely to be vaccine hesitant than those who tested negative by PCR/serology.^12^ This may reflect HCWs with evidence of previous SARS-CoV-2 infection feeling they have derived sufficient immunological protection against COVID-19 via natural infection and will therefore derive limited benefit from vaccination. Whilst this is likely to be true in a short period following the infective episode, over time, reinfection is possible. Population level data from Denmark indicate that infection with SARS-CoV-2 offers 80·5% protection against reinfection, dropping to 47·1% in those over 65.^47^ Furthermore, SARS-CoV-2 neutralising antibody dynamics in those recovered from COVID-19 have been shown to vary widely,^48^ and protective immunity to related seasonal coronaviruses is known to be short-lasting.^49,50^ Therefore, HCWs with evidence of previous COVID-19 (particularly those who were infected many months previously) represent important targets for vaccination, and publicising this message in communications aimed at HCWs may improve uptake in this group.

This is the largest study of SARS-CoV-2 vaccine attitudes in a multi-ethnic sample of UK HCWs at the start of a vaccine roll-out. The combination of quantitative and qualitative data provides an in-depth understanding of hesitancy among different ethnic groups. Despite these strengths, our study also has a number of limitations including the potential for self-selection bias and the low number of ancillary staff in the sample. Due to the rapidly evolving nature of the vaccination programme, questions relating to vaccination were changed midway through the baseline questionnaire rollout which could have impacted on outcome, although we have controlled for questionnaire version in the multivariable analysis. The relevant sections of the baseline questionnaire were not designed to capture actual vaccine uptake as an outcome but rather attitudes towards vaccination and thus we cannot determine whether access to vaccination could be a driver in vaccine hesitancy in our sample, however this will be captured in follow-up questionnaires.

In summary, we have identified key predictors of SARS-CoV-2 vaccine hesitancy in HCWs and demonstrate clear ethnic differences in hesitancy levels. Importantly, we have established drivers behind vaccine hesitancy in HCWs, which include belief in COVID-19 conspiracies and mistrust (of vaccines in general, in SARS-CoV-2 vaccines specifically, in healthcare systems and research) and suggest that these factors may account for some of the observed ethnic differences in hesitancy. Strategies to improve vaccine confidence are urgently required to prevent these ethnic disparities from widening. Such strategies may include building trust and involvement/engagement of ethnic minority HCWs in the vaccination rollout, promoting vaccination and overcoming misinformation utilising trusted networks in ethnic minority communities.

## Data Availability

To access data or samples produced by the UK-REACH study, the working group representative must first submit a request to the Core Management Group by contacting the UK-REACH Project Manager in the first instance. For ancillary studies outside of the core deliverables, the Steering Committee will make final decisions once they have been approved by the Core Management Group. Decisions on granting the access to data/materials will be made within eight weeks.

Third party requests from outside the Project will require explicit approval of the Steering Committee once approved by the Core Management Group.

Note that should there be significant numbers of requests to access data and/or samples then a separate Data Access Committee will be convened to appraise requests in the first instance.

https://www.uk-reach.org/data-dictionary

## Acknowledgements

We would like to thank all the healthcare workers who took part in this study when the NHS was under immense pressure.

We wish to acknowledge the Professional Expert Panel group and the Steering and Advisory Group (see the cohort study protocol^33^ for details), and SERCO, as well as the following people for their support in setting up the study from the regulatory bodies: Kerrin Clapton and Andrew Ledgard (General Medical Council), Caroline Kenny (Nursing and Midwifery Council), David Teeman and Lisa Bainbridge (General Dental Council), My Phan and John Tse (General Pharmaceutical Council), Angharad Jones (General Optical Council), Katherine Timms and Charlotte Rogers (The Health and Care Professions Council) and Mark Neale (Pharmaceutical Society of Northern Ireland).

We would also like to acknowledge the following trusts and sites who recruited participants to the study: Nottinghamshire Healthcare NHS Foundation Trust, University Hospitals Leicester, Lancashire Teaching Hospitals NHS Foundation Trust, Northumbria Healthcare, Berkshire Healthcare, Derbyshire Healthcare NHS Foundation Trust, South Tees NHS Foundation Trust, Birmingham and Solihull NHS Foundation Trust, Affinity Care, Royal Brompton and Harefield, Sheffield Teaching Hospitals, St George’s Hospital, Yeovil District Hospital, Lewisham and Greenwich NHS Trust, Black Country Community Healthcare NHS Foundation Trust, Sussex Community NHS Foundation Trust, South Central Ambulance Service, University Hospitals Coventry and Warwickshire, University Hospitals Southampton NHS Foundation Trust, London Ambulance Trust, Royal Free, Birmingham Community Healthcare NHS Foundation Trust, Central London Community Healthcare, Chesterfield Royal Hospital, Bridgewater Community Healthcare, Northern Borders, County Durham and Darlington Foundation Trust, Walsall Healthcare NHS Trust.

## Author contributions

MP conceived of the idea and led the application for funding with input from MDT, KK, ICM, KW, Robert Free (RF), LBN, SC, Keith R Abrams, LJG, ALG and CJ. The survey was designed by KW, MP, ICM, CMel, CJ, ALG, LBN, RF and CAM. Online consent and survey tools were developed by LB. KW, CAM, ICM, and LBN wrote the first draft of the manuscript with input from MP and all co-authors. All authors approved the submitted manuscript.

## Funding

UK-REACH is supported by a grant from the MRC-UK Research and Innovation (MR/V027549/1) and the Department of Health and Social Care through the National Institute for Health Research (NIHR) rapid response panel to tackle COVID-19.

Core funding was also provided by □NIHR Biomedical Research Centres.

KW is funded through an NIHR Career Development Fellowship (CDF-2017-10-008). CAM is an NIHR Academic Clinical Fellow (ACF-2018-11-004).

LBN is supported by an Academy of Medical Sciences Springboard Award (SBF005\1047).

ALG was funded by internal fellowships at the University of Leicester from the Wellcome Trust Institutional Strategic Support Fund (204801/Z/16/Z) and the BHF Accelerator Award (AA/18/3/34220).

MDT holds a Wellcome Trust Investigator Award (WT 202849/Z/16/Z) and an NIHR Senior Investigator Award.

KK is supported by the National Institute for Health Research (NIHR) Applied Research Collaboration East Midlands (ARC EM).

KK and MP are supported by the NIHR Leicester Biomedical Research Centre (BRC). MP is supported by a NIHR Development and Skills Enhancement Award.

This work is carried out with the support of BREATHE - The Health Data Research Hub for Respiratory Health [MC_PC_19004] in partnership with SAIL Databank. BREATHE is funded through the UK Research and Innovation Industrial Strategy Challenge Fund and delivered through Health Data Research UK.

## Disclaimers

The views expressed in the publication are those of the author(s) and not necessarily those of the National Health Service (NHS), the NIHR or the Department of Health and Social Care. This research was funded in whole, or in part, by the Wellcome Trust [WT204801/Z/16/Z and WT 202849/Z/16/Z]. For the purpose of open access, the author has applied a CC BY public copyright licence to any Author Accepted Manuscript version arising from this submission.

## Competing interests

KK is Director of the University of Leicester Centre for Black Minority Ethnic Health, Trustee of the South Asian Health Foundation, Chair of the Ethnicity Subgroup of the UK Government Scientific Advisory Group for Emergencies (SAGE) and Member of Independent SAGE. SC is Deputy Medical Director of the General Medical Council, UK Honorary Professor, University of Leicester.

## Footnotes

1 The ‘White Other’ group includes the ‘White Other’ and ‘White Gypsy and Irish Traveller’ categories, because there were very small numbers in the latter group.

## Notes

### Clinical Trial

ISRCTN11811602

### Clinical Protocols

https://www.medrxiv.org/content/10.1101/2021.02.23.21251975v1

https://www.medrxiv.org/content/10.1101/2021.03.03.21252737v1.article-info

### Author Declarations

The study was approved by the Health Research Authority (Brighton and Sussex Research Ethics Committee; ethics reference: 20/HRA/4718). All participants gave written informed consent.

### Summary of Updates

In-text references to tables and figures. Updated data sharing statement. Correction of male/female reversal in Table 2.

## References

1. Polack FP, Thomas SJ, Kitchin N, et al. Safety and Efficacy of the BNT162b2 mRNA Covid-19 Vaccine. New England Journal of Medicine 2020; 383(27): 2603–15.

2. Voysey M, Clemens SAC, Madhi SA, et al. Safety and efficacy of the ChAdOx1 nCoV-19 vaccine (AZD1222) against SARS-CoV-2: an interim analysis of four randomised controlled trials in Brazil, South Africa, and the UK. The Lancet 2021; 397(10269): 99–111.

3. Aran D. Estimating real-world COVID-19 vaccine effectiveness in Israel using aggregated counts. medRxiv 2021: 2021·02·05·21251139.

4. Dror AA, Eisenbach N, Taiber S, et al. Vaccine hesitancy: the next challenge in the fight against COVID-19. European Journal of Epidemiology 2020; 35(8): 775–9.

5. Grech V, Gauci C, Agius S. Vaccine hesitancy among Maltese healthcare workers toward influenza and novel COVID-19 vaccination. Early Human Development 2020: 105213.

6. Gagneux-Brunon A, Detoc M, Bruel S, et al. Intention to get vaccinations against COVID-19 in French healthcare workers during the first pandemic wave: a cross-sectional survey. Journal of Hospital Infection 2021; 108: 168–73.

7. Wang K, Wong ELY, Ho KF, et al. Intention of nurses to accept coronavirus disease 2019 vaccination and change of intention to accept seasonal influenza vaccination during the coronavirus disease 2019 pandemic: A cross-sectional survey. Vaccine 2020; 38(45): 7049–56.

8. Sallam M. COVID-19 vaccine hesitancy worldwide: a systematic review of vaccine acceptance rates. medRxiv 2021: 2020·12·28·20248950.

9. Head KJ, Kasting ML, Sturm LA, Hartsock JA, Zimet GD. A National Survey Assessing SARS-CoV-2 Vaccination Intentions: Implications for Future Public Health Communication Efforts. Science Communication 2020; 42(5): 698–723.

10. Prematunge C, Corace K, McCarthy A, Nair RC, Pugsley R, Garber G. Factors influencing pandemic influenza vaccination of healthcare workers—A systematic review. Vaccine 2012; 30(32): 4733–43.

11. Hollmeyer HG, Hayden F, Poland G, Buchholz U. Influenza vaccination of health care workers in hospitals—A review of studies on attitudes and predictors. Vaccine 2009; 27(30): 3935–44.

12. Martin CA, Marshall C, Patel P, et al. Association of demographic and occupational factors with SARS-CoV-2 vaccine uptake in a multi-ethnic UK healthcare workforce: a rapid real-world analysis. medRxiv 2021: 2021·02·11·21251548.

13. Wouters OJ, Shadlen KC, Salcher-Konrad M, et al. Challenges in ensuring global access to COVID-19 vaccines: production, affordability, allocation, and deployment. The Lancet 2021; 397(10278): 9.

14. Hanif W, Ali SN, Patel K, Khunti K. Cultural competence in covid-19 vaccine rollout. BMJ 2020; 371: m4845.

15. Scientific Advisory Group for Emergencies (SAGE): Ethnicity sub-group. Factors influencing COVID-19 vaccine uptake among minority ethnic groups, 2021.

16. Robertson E, Reeve KS, Niedzwiedz CL, et al. Predictors of COVID-19 vaccine hesitancy in the UK Household Longitudinal Study. medRxiv 2021: 2020·12·27·20248899.

17. Paul E, Steptoe A, Fancourt D. Attitudes towards vaccines and intention to vaccinate against COVID-19: Implications for public health communications. The Lancet Regional Health - Europe 2021; 1.

18. Sherman SM, Smith LE, Sim J, et al. COVID-19 vaccination intention in the UK: results from the COVID-19 vaccination acceptability study (CoVAccS), a nationally representative cross-sectional survey. Human Vaccines & Immunotherapeutics 2020: 1–10.

19. National Health Service. NHS Workforce. 2021. https://www.ethnicity-factsfigures.service.gov.uk/workforce-and-business/workforce-diversity/nhs-workforce/latest (accessed 11th February 2021.

20. Robinson E, Jones A, Lesser I, Daly M. International estimates of intended uptake and refusal of COVID-19 vaccines: A rapid systematic review and meta-analysis of large nationally representative samples. Vaccine 2021.

21. Paul E, Steptoe A, Fancourt D. Anti-vaccine attitudes and risk factors for not agreeing to vaccination against COVID-19 amongst 32,361 UK adults: Implications for public health communications. medRxiv 2020: 2020·10·21·20216218.

22. Gadoth A, Halbrook M, Martin-Blais R, et al. Cross-sectional Assessment of COVID-19 Vaccine Acceptance Among Health Care Workers in Los Angeles. Annals of Internal Medicine 2021.

23. Amin DP, Palter JS. COVID-19 vaccination hesitancy among healthcare personnel in the emergency department deserves continued attention. The American Journal of Emergency Medicine 2021.

24. Sze S, Pan D, Nevill CR, et al. Ethnicity and clinical outcomes in COVID-19: A systematic review and meta-analysis. EClinicalMedicine 2020; 29-30: 100630.

25. Pan D, Sze S, Minhas JS, et al. The impact of ethnicity on clinical outcomes in COVID-19: A systematic review. EClinicalMedicine 2020; 23.

26. MacDonald NE. Vaccine hesitancy: Definition, scope and determinants. Vaccine 2015; 33(34): 4161–4.

27. Abbas M, Robalo Nunes T, Martischang R, et al. Nosocomial transmission and outbreaks of coronavirus disease 2019: the need to protect both patients and healthcare workers. Antimicrobial Resistance & Infection Control 2021; 10(1): 7.

28. Paterson P, Meurice F, Stanberry LR, Glismann S, Rosenthal SL, Larson HJ. Vaccine hesitancy and healthcare providers. Vaccine 2016; 34(52): 6700–6.

29. Larson HJ, Jarrett C, Eckersberger E, Smith DMD, Paterson P. Understanding vaccine hesitancy around vaccines and vaccination from a global perspective: A systematic review of published literature, 2007–2012. Vaccine 2014; 32(19): 2150–9.

30. The Sage Working Group on Vaccine Hesitancy. Report of the Sage Working Group on Vaccine Hesitancy, 2014.

31. Barry M, Temsah M-H, Aljamaan F, et al. COVID-19 vaccine uptake among healthcare workers in the fourth country to authorize BNT162b2 during the first month of rollout. medRxiv 2021: 2021·01·29·21250749.

32. Shekhar R, Sheikh AB, Upadhyay S, et al. COVID-19 Vaccine Acceptance among Health Care Workers in the United States. Vaccines 2021; 9(2): 119.

33. Woolf K, Melbourne C, Bryant L, et al. The United Kingdom Research study into Ethnicity And COVID-19 outcomes in Healthcare workers (UK-REACH): Protocol for a prospective longitudinal cohort study of healthcare and ancillary workers in UK healthcare settings. medRxiv 2021: 2021·02·23·21251975.

34. Gogoi M, Reed-Berendt R, Al-Oraibi A, et al. Ethnicity and COVID-19 outcomes among healthcare workers in the United Kingdom: UK-REACH ethico-legal research, qualitative research on healthcare workers’ experiences, and stakeholder engagement protocol. medRxiv 2021: 2021·03·03·21252737.

35. Ritchie J, Spencer L. Qualitative Data Analysis for Applied Policy Research. In: Huberman AM, Miles MB, editors. The Qualitative Researcher’s Companion. Thousand Oaks, California: SAGE Publications, Inc.; 2002.

36. Sunstein C AE, Kim M, Carrasco M, Chadborn T, Gauri V, et al. Behavioural considerations for acceptance and uptake of COVID-19 vaccines. World Health Organisation Technical Advisory Group on Behavioural Insights and Sciences for Health Meeting Report 15th October 2020. Geneva: World Health Organisation, 2020.

37. Wang J, Feng Y, Hou Z, et al. Willingness to receive SARS-CoV-2 vaccine among healthcare workers in public institutions of Zhejiang Province, China. Human Vaccines & Immunotherapeutics 2021: 1–8.

38. Ledda C, Costantino C, Cuccia M, Maltezou HC, Rapisarda V. Attitudes of Healthcare Personnel towards Vaccinations before and during the COVID-19 Pandemic. International Journal of Environmental Research and Public Health 2021; 18(5).

39. Qattan AMN, Alshareef N, Alsharqi O, Al Rahahleh N, Chirwa GC, Al-Hanawi MK. Acceptability of a COVID-19 Vaccine Among Healthcare Workers in the Kingdom of Saudi Arabia. Front Med (Lausanne) 2021; 8: 644300.

40. Kwok KO, Li K-K, Wei WI, Tang A, Wong SYS, Lee SS. Influenza vaccine uptake, COVID-19 vaccination intention and vaccine hesitancy among nurses: A survey. International Journal of Nursing Studies 2021; 114: 103854.

41. Kuter BJ, Browne S, Momplaisir FM, et al. Perspectives on the receipt of a COVID-19 vaccine: A survey of employees in two large hospitals in Philadelphia. Vaccine 2021; 39(12): 1693–700.

42. Unroe KT, Evans R, Weaver L, Rusyniak D, Blackburn J. Willingness of Long-Term Care Staff to Receive a COVID-19 Vaccine: A Single State Survey. Journal of the American Geriatrics Society 2021; 69(3): 593–9.

43. Campbell L. Care home workers in England face mandatory Covid jabs under plans. The Guardian. 2021.

44. Paterlini M. Covid-19: Italy makes vaccination mandatory for healthcare workers. BMJ 2021; 373: 905.

45. Khunti K, Kamal A, Pareek A, Griffiths A. Should vaccination for healthcare workers be mandatory? Journal of the Royal Society of Medicine Forthcoming.

46. Allington D, Duffy B, Moxham-Hall V, McAndrew S, Murkin G. Coronavirus conspiracies and views of vaccination. 2021. https://www.kcl.ac.uk/policy-institute/assets/coronavirus-conspiracies-and-views-of-vaccination.pdf (accessed 18th April 2021).

47. Hansen CH, Michlmayr D, Gubbels SM, Mølbak K, Ethelberg S. Assessment of protection against reinfection with SARS-CoV-2 among 4 million PCR-tested individuals in Denmark in 2020: a population-level observational study. The Lancet 2021; 397(10280): 1204–12.

48. Chia WN, Zhu F, Ong SWX, et al. Dynamics of SARS-CoV-2 neutralising antibody responses and duration of immunity: a longitudinal study. The Lancet Microbe.

49. Edridge AWD, Kaczorowska J, Hoste ACR, et al. Seasonal coronavirus protective immunity is short-lasting. Nature Medicine 2020; 26(11): 1691–3.

50. Boyton RJ, Altmann DM. Risk of SARS-CoV-2 reinfection after natural infection. The Lancet 2021; 397(10280): 1161–3.

