## Supplementary information for "Ethnic differences in SARS-CoV-2 vaccine hesitancy in United Kingdom healthcare workers: Results from the UK-REACH prospective nationwide cohort study"

**Supplementary Figure 1: Diagram demonstrating how the primary outcome measure of Vaccine Hesitancy was derived from two vaccine questions (VQ1 administered from 4^th^ to 20^th^ December 2020, and VQ2 administered from 21^st^ December 2020 onwards).** PNTA=prefer not to answer. Orange boxes indicate responses coded as Hesitant, green boxes indicate responses coded as Not Hesitant in the primary outcome measure of Vaccine Hesitancy.

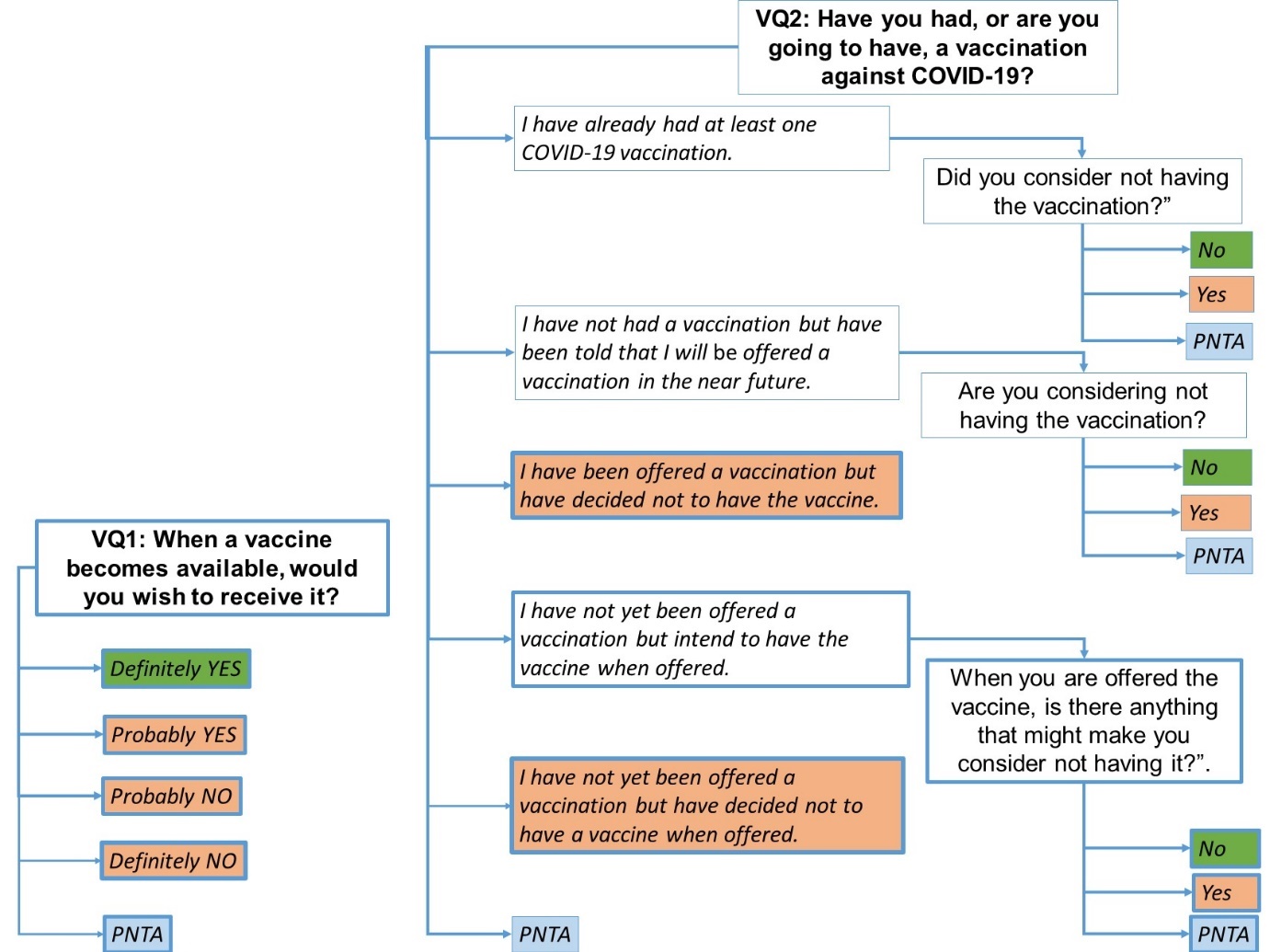

**Supplementary Table 1: Variables included in the imputation model.** Variables are grouped by questionnaire section, which were presented in the order described. Further details on variables can be found in the Data Dictionary*^[1]^*. Raw items refer to individual variables in the survey software programme RedCap, and in many cases involve logic chains so that not all participants see all questions, or have very many raw items due to the multiple checkbox format. Derived variables sometimes have the same information in different formats (e.g. age in years, age in decadal groups, etc). Imputation was on the basis of 214 variables. 4∙58% of data points were missing (113,751/2,478,976), with a median of 7 per participant (mean: 9∙82, SD 8∙53, 95% range = 0 to 31); 927/11,584 (8∙0%) of participants had complete data. The imputation model included all derived questionnaire variables. Collinear variables were omitted.

| *Section* | *Content area* | *Raw* | *Derived* |
| --- | --- | --- | --- |
| 0: Registration | Date of consent and completion; questionnaire completed; regulator; age in years; Indices of Multiple Deprivation (IM), derived from registration postcode | 15 | 27 |
| 1: Demographics | Sex; gender; relationship status | 4 | 3 |
| 2: Job | Job/role; profession; specialty; working during first lockdown; sector; location; UK region; grade; NHS band; work areas now and in lockdown; night work; contact/communication with patients with and without COVID; travel to work; access to PPE (Personal Protective Equipment); aerosol generating procedures; risk assessment; concerns about unsafe practice; role change during pandemic | 195 | 63 |
| 3: Ethnicity, culture and religion; home and family life | Ethnicity (ONS 18 groups); country of birth; nationality(s); ethnicity etc of partner, mother and father; grandparents born in UK; languages spoken at home as a child; age of learning English; Religion (ONS categories); importance of religion and attending place of worship; importance of cultural and ethnic identity; country of primary qualification; level of education of self, mother and father; work colleagues of same ethnicity as oneself | 107 | 104 |
| 4: Home and family life | Support bubble; childcare bubble; people in household (age, sex, relationship); travel to work of household; current accommodation; rooms; outdoor space | 46 | 30 |
| 5: Friends and social network | People talked with remotely/face-to-face/ with physical contact; friends of same ethnicity as self | 4 | 4 |
| 6: Harassment and discrimination | Everyday Discrimination Scale; discrimination at work; complained about discrimination at work | 50 | 5 |
| 7: Health | Height and weight; smoking and vaping; alcohol use; exercise activities; walking pace; lifestyle changes in lockdown; hospital and GP visits; flu vaccination; shielding; medications and supplements; comorbidities; EQ-5D health scales; anxiety (GAD2); depression (PHQ2); PTSD (PCL-C); financial worries; loneliness (UCLA); life satisfaction | 74 | 66 |
| 8: COVID-19 | COVID-19 contacts; behavior changes during lockdown; response to lockdown; swan and antibody tests; symptoms in past two weeks; had COVID-19 infection; concerns about getting COVID-19, spreading COVID-19; personally know people dying from COVID-19; sources of information about COVID-19; perceived personal risks of catching COVID-19; vaccination attitudes (different questions in December and January onwards); been vaccinated; attitudes to vaccination (VAX scale); optimism about speed of vaccination programme; effects of COVID-19 on society; COVID-19 knowledge and COVID-19 conspiracy beliefs; | 237 | 87 |
| 9: Approach to Life | Big Five personality scale; Locus of control scale; Fatalism scale; Need for Cognitive Closure scale; approach to risk-taking; Work-place climate (pre-COVID-19); burnout (pre-COVID-19) | 43 | 18 |
| 10: Final questions | Open-ended questions (Why have people from ethnic minorities been more severely affected by COVID-19? How will society change as a result of COVID-19? How do you see your own future changing because of COVID-19?); Was the questionnaire too long or too short?; How useful will the questionnaire be for understanding COVID-19 in ethnic minorities? | 5 | 10 |
| *Total* |  | *780* | *417* |

**Supplementary Table 2: List of variables included in the study.** Variables grouped into categories measuring sociodemographic factors, vaccine access, trust in vaccines or those giving the vaccines, perceived risk of COVID-19, and psychological factors.

| **Variable** | **Levels** | **Variable category** |
| --- | --- | --- |
| Gender | Binary | Sociodemographic |
| Age (years) | Continuous | Sociodemographic |
| Ethnicity (17 UK Office for National Statistics categories) | Dummy variables, reference category: White British | Sociodemographic |
| Country of birth | Binary (UK, not UK) | Sociodemographic |
| Religion (Christian, Buddhist, Hindu, Jewish, Muslim, Sikh, Other, None) | Dummy variables, ref: None | Sociodemographic |
| Index of multiple deprivation decile | Ordinal | Sociodemographic |
| Job category (Nurse, Allied Health Professional, Dental, Administration or other, Doctor) | Dummy variables, ref: doctor | Access |
| Job sector | Binary (NHS, not NHS) | Access |
| Job location | 3 levels: (Hospital, General Practice, Community | Access |
| Influenza Vaccine 2019/2020 | Binary (vaccinated, not vaccinated) | Trust/Access |
| COVID conspiracy beliefs | Continuous: 1 (all definitely true) to 24 (all definitely false). Derived from sum of scores on six items rated on a four-point scale. | Trust |
| Pro-vaccine attitudes | Continuous | Trust |
| Discriminated against at work on the basis of ethnicity, nationality and/or religion | Binary (discriminated against, not discriminated against) | Trust |
| Trust in employer: secure raising concerns about poor clinical practice | Continuous/ordinal (1 to 5) | Trust |
| Trust in employer: confident concerns would be addressed | Continuous/ordinal (1 to 5) | Trust |
| Fatalism | Continuous (5 to 35) | Psychological/ Perceived risk |
| Personality: Agreeable | Continuous | Psychological |
| Personality: Conscientious | Continuous | Psychological |
| Personality: Neuroticism | Continuous | Psychological |
| Personality: Openness | Continuous | Psychological |
| Personality: Extraversion | Continuous | Psychological |
| Pregnant | Binary (pregnant, not pregnant) | Perceived risk |
| Number of non-psychiatric comorbidities | Ordinal: 0, 1, 2+ | Perceived risk |
| Body Mass Index | Continuous/Ordinal | Perceived risk |
| COVID test positivity | Three levels: positive, not tested, tested negative | Perceived risk |
| Concern about unknowingly spreading COVID | Binary (not concerned, very or quite concerned) | Perceived risk |
| Perception of COVID risk to self | Continuous (0 optimistic to 100 pessimistic) | Perceived risk |
| Number of COVID patients seen per week face to face | Binary (exposed, not exposed). | Perceived risk |
| COVID information source: Official sources | Continuous: mean of six information sources: (NHS website + Government NHS website+ UK Government website+ my own GP + Local Council + Welsh Scottish NI Government websites)/6. | Information source |
| COVID information source: Personal contacts or social media | Continuous: mean of five information sources: (Friends or family+ Social media+ colleagues + twitter + employer)/5. | Information source |
| COVID information source: Mainstream media | Continuous: mean of three information sources (TV+ Radio+ newspaper)/3. | Information source |
| COVID information source: Scientific sources | Continuous, although mean of three information sources (Science Journals+ Other websites + WHO website)/3. | Information source |
| Vaccine question answered | Binary (VQ1, VQ2) |  |

**Supplementary Table 3: Reasons for vaccine hesitancy by ethnicity, ordered from most frequently selected to least frequently selected.** Question asked only of those who answered VQ2 (missing n=1293). Participants could select as many reasons as they wanted. ^*^ethnic difference significant at p<=0∙001. Within the “other reason” category, freetext comments category mostly related to pregnancy, breastfeeding or fertility (n=162; 69%); 17 (7%) related to having had COVID, and either being concerned about the effects on persistent symptomatic (“long”) COVID or preferring “natural” immunity; 10 (4%) related to concerns about the decision by the UK government to delay the second dose to 12 weeks; and nine (4%) were concerns that the vaccine has animal, foetal or blood products in it. The remainder (n=62; 26%) related to concerns about side effects, perceived lack of scientific testing or lack of trust in the scientific establishment and/or government, being unwell when offered the vaccine, or being in a vaccine trial.

| **Reason for hesitancy** | **n (% of total n=2792);** | **White n (%)** | **Asian n (%)** | **Black n (%)** | **Mixed n (%)** | **Other n (%)** |
| --- | --- | --- | --- | --- | --- | --- |
| Concerned about the safety or potential side-effects^*^ | 946 (63∙1) | 546 (59∙3) | 225 (71∙4) | 93 (68∙9) | 32 (69∙2) | 49(63∙6) |
| Prefer to wait until many other people have received a COVID-19 vaccine^*^ | 417 (27∙8) | 221 (24∙0) | 104 (33∙0) | 50 (37∙0) | 16 (34∙8) | 26 (33∙8) |
| Would rather the vaccine were used for other people who need it more than I do | 298 (19∙9) | 189 (20∙5) | 61 (19∙4) | 19 (14∙1) | 10 (21∙7) | 18 (23∙4) |
| Not convinced that COVID-19 vaccines will be effective | 285 (19∙0) | 173 (18∙8) | 60 (19∙0) | 26 (19∙3) | 10 (21∙7) | 10 (21∙7) |
| Vaccines may not have been tested thoroughly in all ethnic groups^*^ | 277 (18∙5) | 63 (6∙8) | 109 (34∙6) | 63 (46∙7) | 16 (34∙8) | 26 (33∙8) |
| Other reason | 264 (17∙6) | 174 (18∙9) | 49 (15∙6) | 18 (13∙3) | 5 (10∙9) | 16 (20∙8) |
| Allergies/needle-phobia/immuno-compromised/other clinical reason | 197 (13∙1) | 136 (14∙8) | 29 (9∙2) | 11 (8∙1) | 7 (15∙2) | 13 (16∙9) |
| Had COVID-19 and therefore do not feel I need the vaccine | 138 (9∙2) | 74 (8∙0) | 31 (9∙8) | 20 (14∙8) | 5 (10∙9) | 7 (9∙1) |
| Do not feel that I personally am at risk from COVID-19 | 134 (8∙9) | 88 (9∙6) | 18 (5∙7) | 14 (10∙4) | 5 (10∙9) | 8 (10∙4) |
| Prefer one of the other COVID-19 vaccines that are being developed | 126 (8∙4) | 80 (8∙7) | 20 (6∙3) | 13 (9∙6) | <5 (<2) | 11 (14∙3) |
| Do not believe in vaccinations in general | 19 (1∙3) | 11 (1∙2) | <5 (<2) | <5 (<2) | <5 (<2) | <5 (<2) |
| Taking part in a clinical trial of a COVID-19 vaccine | 9 (0∙6) | 7 (0∙8) | <5 (<2) | <5 (<2) | <5 (<2) | <5 (<2) |

**Supplementary Table 4: Predictors of vaccine hesitancy stratified by ethnicity**

| **Variable** | **Ethnicity** | | | | |
| --- | --- | --- | --- | --- | --- |
|  | **White** | **Asian** | **Black** | **Mixed** | **Other** |
| **TRUST VARIABLES** | | | | | |
| **Belief in COVID-19 ‘conspiracies’ score, med (IQR)**  (min 6 [does not believe] - max 24 [strongly believes]) | 8 (7 - 10) | 9 (8 - 11)** | 10 (8 - 12)** | 8 (7 - 10) | 10 (8 - 12)** |
| **Pro-vaccine score**, **med (IQR)**  (min 4 [anti-vaccination] - max 20 [pro-vaccination]) | 16 (14 - 17) | 15 (13 - 17)** | 15 (13 - 16)** | 16 (14 - 17) | 15 (13 - 17)** |
| **Influenza vaccination status 2019 - 2020**, **n(%)**  Vaccinated  Unvaccinated | 5747 (72∙3%)  2206 (27∙7%) | 1530 (70∙6%)  637 (29∙4%) | 309 (66∙2%)  158 (33∙8%)* | 351 (74∙8%)  118 (25∙2%) | 181 (74∙5%)  62 (25∙5%) |
| **Trust in employer to deal with a concern about unsafe clinical practice, n(%)**  1 (does not trust employer)  2  3  4  5 (trusts employer) | 224 (3∙0%)  634 (8∙6%)  1148 (15∙5%)  2841 (38∙4%)  2552 (34∙5%) | 82 (4∙0%)  200 (9∙9%)  433 (21∙3%)  769 (37∙9%)  547 (26∙9%)** | 20 (4∙7%)  30 (7∙1%)  79 (18∙6%)  167 (39∙4%)  128 (30∙2%) | 14 (3∙2%)  38 (8∙6%)  80 (18∙1%)  180 (40∙6%)  131 (29∙6%) | 10 (4∙4%)  27 (11∙8%)  41 (17∙9%)  88 (38∙4%)  63 (27∙5%)* |
| **Discrimination at work on the basis of ethnicity, nationality or religion, n(%)**  Has not experienced discrimination  Has experienced discrimination | 5553 (75∙1%)  1839 (24∙9%) | 1187 (58∙7%)  837 (41∙4%)** | 208 (47∙8%)  227 (52∙2%)** | 284 (64∙1%)  159 (35∙9%)** | 133 (59∙4%)  91 (40∙6%)** |
| **RISK VARIABLES** | | | | | |
| **Previous laboratory evidence of**  **COVID-19 (PCR or serology), n(%)**  Never tested  Tested negative  Tested positive | 1275 (16∙1%)  5117 (64∙4%)  1549 (19∙5%) | 339 (15∙6%)  1364 (62∙8%)  469 (21∙6%) | 80 (17∙1%)  280 (59∙7%)  109 (23∙2%) | 77 (16∙4%)  305 (64∙8%)  89 (18∙9%) | 25 (10∙3%)  163 (66∙8%)  56 (23∙0%) |
| **Number of comorbidities, n(%)**  0  1  ≥2 | 5495 (71∙6%)  1712 (22∙3%)  468 (6∙1%) | 1444 (69∙3%)  487 (23∙4%)  153 (7∙3%) | 294 (65∙5%)  120 (26∙7%)  35 (7∙8%) | 313 (69∙3%)  105 (23∙2%)  34 (7∙5%) | 155 (67∙7%)  60 (26∙2%)  14 (6∙1%) |
| **Perceived risk of hospitalisation with COVID-19, med, (IQR)**  (100 point scale) | 15 (5 - 40) | 25 (10 - 50)** | 20 (5 - 50) | 20 (5 - 45) | 20 (8 - 50) |
| **Concerned about unknowingly spreading COVID-19, n(%)**  Not concerned  Concerned | 3895 (48∙9%)  4078 (51∙2%) | 1127 (52∙0%)  1042 (48∙0%) | 266 (57∙6%)  196 (42∙4%)** | 220 (47∙1%)  247 (52∙9%) | 133 (55∙0%)  109 (45∙0%) |
| **Exposed to COVID-19 patients at work, n(%)**  Unexposed  Exposed | 5226 (69∙6%)  2278 (30∙4%) | 1173 (57∙4%)  869 (42∙6%)** | 240 (55∙7%)  191 (44∙3%)** | 290 (65∙0%)  156 (35∙0%) | 118 (50∙6%)  115 (49∙4%)** |

**p<0∙001, *p<0∙01 compared to White using chi-squared test or Wilcoxon rank-sum test as appropriate

**Supplementary Tables 5-8: Quotes on drivers of vaccine hesitancy (trust, perceived risk, health information and messaging) and improving delivery.**

**Table S5: Trust**

Pleased to be prioritised:

I mean I’m privileged to have had it so early. I think about certain parts of the world where you’re not going to even get access to it this year. So yeah, I feel very lucky and privileged to live in a country where I was able to access the vaccine.

[Participant 1, Clinical Pharmacist]

Trust in vaccine science:

Things like MMR, I did get my sons inoculated even though, at the time when they were born there was that whole controversy about the MMR vaccine, but I knew the science was sound. So, for me, the vaccination is positive.

[Participant 6, Estates and Facilities Advisor]

Lack of trust due to speed of development:

I’ve not had the vaccine and I don’t plan to have it…I had concerns it is developed too quick, not the way we were hearing from the news and on the internet…not proper testing is done or it’s not sufficient. So I was hesitant, although I was offered working in the health environment. But I declined.

[Participant 21, Project Manager]

Lack of trust in vaccine:

I am quite worried and mainly about the impact on fertility, you know, and even though studies say there’s no link, there’s no plausible biological mechanism, we haven't had that long term research really to say it’s definitely 100% safe.

[Participant 35, Public Health Registrar]

It’s more like wanting to get the facts right first. As medics you want to know is it efficient? Is it effective to do what it’s said to do? That was my initial fear.

[Participant 28, Consultant ]

Trust in those producing vaccines:

As a […] patient, some of the drugs that we have as Black women, actually it’s having a different effect on us…So I just wanted to know how many people from the Black community, the Asian community, had been involved in the trials.

[Participant 27, Radiographer]

Influence of others on trust in vaccine:

It was very anxiety provoking actually going to have the vaccine. I've got a [child] who’s a doctor, and so [my child] has seen people dying, to be blunt, and basically has taken it and basically said ‘Dad, you’ve got to take it, Mum, you’ve got to take it.’ So I trust [my child], so I decided to take it.

[Participant 34, Consultant Psychologist]

When you see that kind of behaviour you start thinking right, so what’s going on?...You see in the news ‘Oh, the NHS is under pressure and people are dying,’ and suddenly boom, you see…[consultants] sharing drinks from the one straw, sitting together.

[Participant 17, Catering Retail Manager]

Contradiction of personal beliefs and public role:

I haven't been racing to take it, it’s a bit of a contradiction because I know I spend my days trying to improve vaccine uptake. I am still quite worried about it, you know, even though we have to say the official lines about oh it’s safe.

[Participant 35, Public Health Registrar]

**Table S6: Perceived risk**

Perceptions of individual risk:

You’re hearing so much on the news about the wide-ranging complications and although I was in a low-risk group I’m still fearful of getting it and you are exposing yourself going to work regularly when everyone else is told to try and minimise and stay at home as much as possible.

[Participant 5, Doctor]

I think the fact that there are more cases of people who are showing severe signs of the disease in my ethnicity, that’s the first thing. Secondly, I know a couple of friends who’ve got what look like long COVID now…It’s not as easy as ‘Oh, if you’re fit and healthy you’re going to be all right’. I’m not taking that for granted.

[Participant 2, Pharmacist]

Covid-19 is a new virus, there’s huge risk factors associated with that. Some people are absolutely fine - it doesn’t impact them long-term. But other people have had long Covid…I just weighed up the risk…and the risk for having the vaccine slightly won over the risk of Covid.

[Participant 33, Physiotherapist]

Risk to others:

But the main reason I got it was really to protect - it was less to protect myself and more probably to protect my wife who isn't able to have the vaccine at the moment, so it was really to protect her. That was what pushed me to get it.

[Participant 13, Speciality Trainee ]

I didn’t feel like I needed it but I got it because I wanted to show people that I know, people who are afraid of vaccines, who are vaccine hesitant - so my mum, my dad and my family - actually look, I’ve had it, I’m completely fine.

[Participant 2, Pharmacist]

Increased risk of Black, Asian, and Minority Ethnic (BAME) groups and lack of transparency about prioritisation of BAME groups:

I think within six months of the pandemic it was quite clear that the BAME community was more vulnerable to the Covid virus, and you know when the vaccine programme was implemented in the Trust, I could not see why the Trust could not prioritise the BAME community to have the vaccines.

[Participant 41, Consultant]

Spotlight on ethnic minority communities:

I really don’t like that putting together of Covid-19 and ethnicity because as somebody who does research, we have to change some of the terminology that’s used in the NHS and what it gives people the impression is that we’re the spreaders.

[Participant 27, Radiographer]

‘Why are Black doctors not taking the vaccine? Why do Blacks not take the vaccine?’ If you look at the BBC website today, when it talks about the over-80 years of age who are taking the vaccine, Blacks were about 30%. The question they didn’t say is how many, in the population group of over 80 in this country, what percentage are Blacks? So I’m sure the Blacks over 80 in this country make less than 20%...Nobody looks at it…what are the particular reasons why you haven’t had your vaccine. That’s my concern where at this present time, they’re still spinning the story and we are still being victimised about it. So we’ve got to be very careful about that.

[Participant 28, Consultant ]

**Table S7: Health information and messaging**

Trustworthiness and proactively seeking health information:

I think when it came to vaccinations, I tried to get hold of the original research papers myself and read the BMJ a bit more and sort of ignored the news because I wanted to understand it a bit more on a scientific level…so I wanted to read the original papers and make my own mind up.

[Participant 33, Physiotherapist]

Social media:

A lot of people still rely on the wrong messages from WhatsApp and Facebook, a lot of false information.

[Participant 11, Nurse Specialist]

Social media plays a big big role these days. I think most people get their information from social media rather than what the government says…on social media you don’t half the time know what is right and what is wrong. If the same falsehood is being sort of hammered again and again and again, people sort of come to believe that that’s the truth…that misinformation needs to be controlled.

[Participant 41, Consultant]

A really good doctor…I saw the video somewhere uploaded on YouTube or something, who actually explained the reason why the vaccine has been developed that quick…….. and I think that this video hit me and so I said ‘OK, I’m just going to take it’.

[Participant 17, Catering Retail Manager]

Quantity of information:

We do have daily emails about it! But obviously I don't exactly have time to read all the emails very carefully, so I haven't been paying too much attention because I know I wouldn't be having mine any time soon.

[Participant 9, Trainee Scientist]

Inaccessible and complex information:

Probably I will [have the vaccine], but now there’s confusion between this Oxford thing and Pfizer and I don’t know if a third company will come. I’m confused again. I don’t know.

[Participant 21, Project Manager]

Lack of knowledge is the main reason or false rumours running around the community…a multinational community…some of them can't speak [English]…the information which is available in the English, they are not getting access for that. I came across a few patients like that because they don't have any knowledge about how the vaccination’s working…what is going on about vaccination.

[Participant 11, Nurse Specialist]

Stigma around hesitancy and vaccine knowledge:

I feel like there’s a great pressure in the working environment to get the vaccine, especially if you’re in the office and stuff like that, which can make you sometimes feel like - you’re kind of like dehumanised almost and some people feel like you’re maybe desensitised to the situation simply because you don’t want to take the vaccine.

[Participant 18, Management Trainee]

Lack of trust in government honesty around Covid:

I think in this age, you don’t know what to trust. There are all kinds of information coming and I don’t know if the things that I’m reading on the NHS…when they have been claiming that there are only 200 bodies when we know, the inside story, there were much more. Do I believe what they’re telling me about vaccinations now? So there is nothing you can 100% believe yes, that is the thing. If the NHS says something else, I’m not too sure because on other grounds they have lied to me, which I know they have lied to me.

[Participant 21, Project manager]

You had the politicians, then you had the scientists at times conveying very confusing messages, and it became farcical to be quite honest with you, so we had guidance and then the guidance was being changed, we had uproars with regards to consistency. I was hearing lots and lots of things from the point of view of, you know, not to trust the government as to what they were putting out and that there was a lot of spin on it.

[Participant 34, Consultant ]

**Table S8: Improving delivery**

Improving transparency and trust:

Making sure that the information’s available to them about exactly which staff are going to be eligible…hopefully through emails and communication through the Trust we’ll be able to make a decision.

[Participant 5, Medical Doctor]

Equity, access, and opportunity

When the vaccines got rolled out, they gave them to…all the A&E doctors, all the intensive care. It took one of my colleagues to email some people to say, ‘Look, we’re high risk as well, you need to give us vaccines, because we’re seeing people at the front door as well.’ So we got them, and then one of my colleagues went to get a vaccine and was told…‘You don't have a Trust assignment number, we can't give it to you’…It’s surprising how little insight some people have as to the role that junior doctors, trainee doctors play…I think we do get forgotten about sometimes…people worry about money more than they worry about the actual staff that are providing the service. So I would say we were kind of a bit forgotten about more so than some of the lower risk staff categories that were paid by the Trust.

[Participant 13, Geriatric Doctor]

It should really be carefully considered who the higher risk people are.... I think it will be very difficult to identify these people because my experience is that sometimes they’re not necessarily the substantive, regular workers in organisations. I think they’re sometimes bank workers, they’re locum workers, they’re people that work a lot of out of hours shifts, so they work perhaps nights all the time. They’re almost always from ethnic minorities. They’re almost always from larger, multigenerational households, and some of them will often not always have access to the things that the Trust will provide... So I think there’s a really interesting thing here around helping workers that provide a large volume of frontline service, perhaps as locum staff or bank staff and ensuring that they get access to the vaccine early.

[Participant 8, Specialist Registrar]

Mandating vaccination:

Legislation is required to prevent organisations insisting on people being vaccinated and discriminating against those who choose not to have a vaccine.

[Participant 62, Speech and language therapist]

Mandatory vaccinations will only isolate people with conscientious and scientific objections, and cause those who blindly follow the powerful people to become hateful to others.

[Participant 74, Occupational therapist]

I don’t really think it’s practical to just say ‘Ok, every staff member should take a vaccine. You guys have to take the vaccine. Why wouldn’t you want to take the vaccine?’…That doesn’t really do much for the people that have concerns about it.

[Participant 18, Management Trainee]

Outreach through involvement:

It’s a really difficult one from the point of view of persuading our ethnic minority groups, but I think we have to keep doing it and it has to be coming from us, if that makes sense, not from the politicians say but from us. And that’s what I've been doing, you know, I've been promoting it with regards to my work…I'm working with BME [Black and Minority Ethnic] culturally diverse families and I'm saying, ‘Look, you know, you need to be taking this.’

[Participant 34, Consultant ]

I’m considering just doing my vaccine publicly and getting it out in the hospital media and the local media. Not just because I’m Black, you know, there are low-income households saying the same thing.

[Participant 27, Radiographer]

1. [https://www.uk-reach.org/data-dictionary](https://eur01.safelinks.protection.outlook.com/?url=https%3A%2F%2Fwww.uk-reach.org%2Fdata-dictionary&data=04%7C01%7C%7Cd4f50195d8f742418c6408d8d3480a0d%7C1faf88fea9984c5b93c9210a11d9a5c2%7C0%7C0%7C637491653649753313%7CUnknown%7CTWFpbGZsb3d8eyJWIjoiMC4wLjAwMDAiLCJQIjoiV2luMzIiLCJBTiI6Ik1haWwiLCJXVCI6Mn0%3D%7C1000&sdata=oDZFWuPFIzxtWwCl1mrDOqzTagPtClLjkhuS1xHnDv4%3D&reserved=0)
