## Supplementary figures and images for "Ethnic differences in SARS-CoV-2 vaccine hesitancy in United Kingdom healthcare workers: Results from the UK-REACH prospective nationwide cohort study"

### Supplementary Figure 1

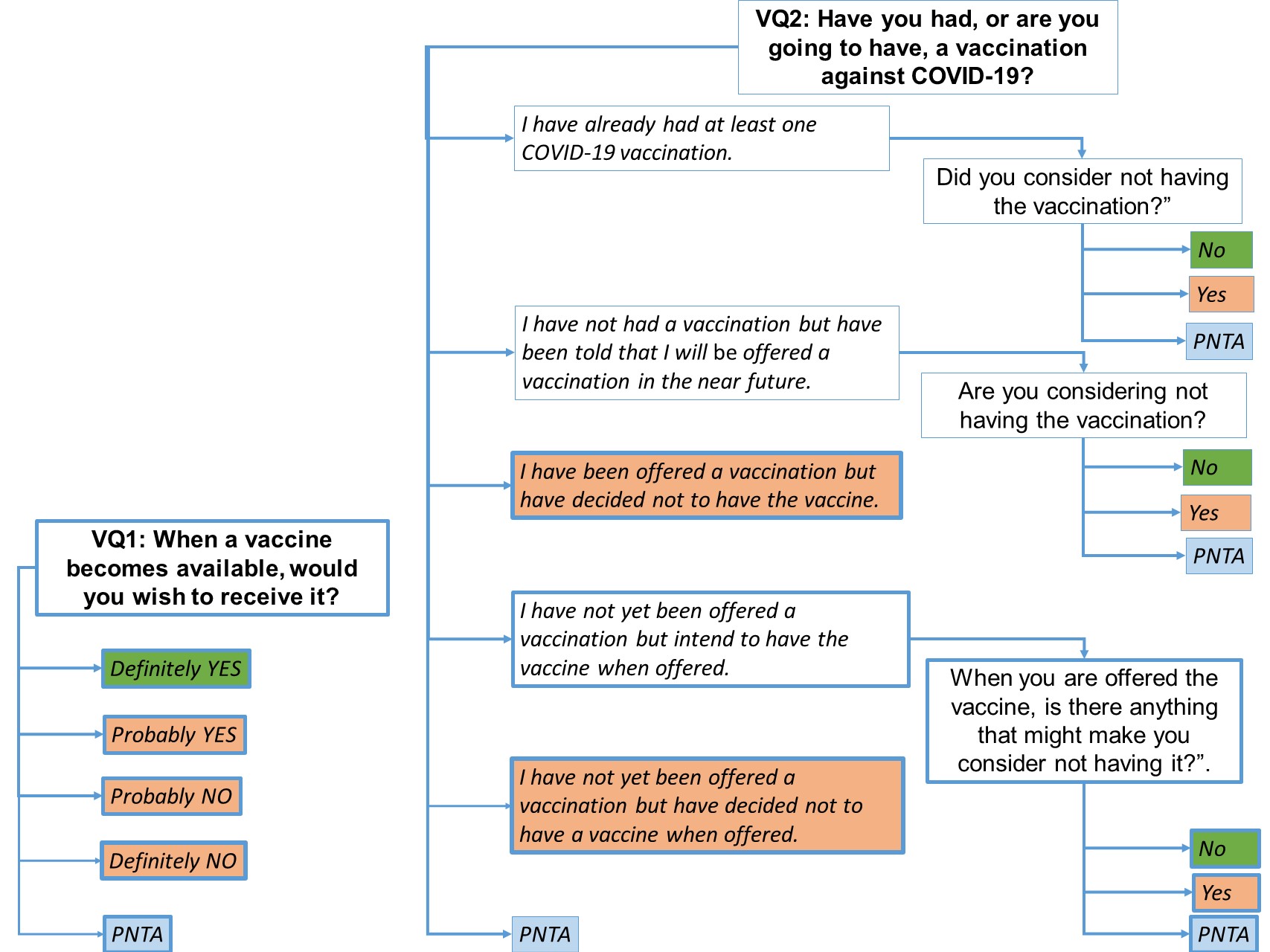
